# Genetic risk factors for postoperative complications after major surgery and shared genetic aetiology with non-postoperative phenotypes

**DOI:** 10.64898/2025.12.11.25342055

**Authors:** Richard A. Armstrong, Paul Yousefi, Ben Gibbison, Golam M Khandaker, Tom R Gaunt

**Author notes:** **Correspondening author:** Richard A Armstrong, University of Bristol, Orchard Lane, Bristol BS1 5DS, UK.

## Abstract

**Background:** Up to 15% of patients worldwide experience serious postoperative complications after major surgery. We explored the genetic basis of five common postoperative complications and whether they share genetic aetiology with their non-postoperative equivalents.

**Methods:** We performed case-control genome-wide association studies (GWAS) using UK Biobank data from >140,000 participants. The primary outcomes were postoperative acute kidney injury (AKI), atrial fibrillation (AF), myocardial infarction (MI), stroke and surgical site infection (SSI). Additionally, we assessed genetic correlation between UK Biobank postoperative outcomes, as well as between these and their non-postoperative (undifferentiated) equivalents from the FinnGen cohort. Finally, we assessed the association between observed postoperative outcomes and polygenic risk scores (PRS) for their non-postoperative equivalents.

**Results:** Among 8,472 postoperative complications in 140,563 eligible participants, three genome-wide significant risk loci for AF were identified on chromosomes 1 (*KCNN3*), 4 (*PITX2*) and 16 (*TRAF7:CASKIN1*). No genome-wide significant loci were found for AKI, MI, stroke or SSI. Strong genetic correlation between postoperative and non-postoperative AF was found (*r_g_* = 0.69, p = 0.001). Increasing PRS quintiles for non-postoperative AF, MI, stroke and chronic kidney disease were associated with increased odds of developing the corresponding postoperative complication.

**Conclusions:** We identified multiple genomic risk loci for postoperative AF, including a putative novel locus on chromosome 16, and demonstrated shared genetic architecture with non-postoperative AF. Although no shared genetic basis was found across different complications, the association between PRS for several non-postoperative phenotypes and their postoperative equivalents suggests that postoperative complications may reflect underlying chronic disease vulnerability with potential implications for targeted risk stratification and postoperative follow-up.

---

Up to 15% of patients worldwide experience serious postoperative complications after major surgery with significant implications for morbidity, mortality and resource utilisation ^1^. These complications represent the interplay between intervention-specific outcomes, the insult of surgery more generally, and underlying vulnerabilities in individuals. They are poorly understood from an aetiological perspective and have few clinically practical predictors to guide preventive treatment ^2,3^.

There is increasing recognition that genetic influences can impact postoperative outcomes and that perioperative care needs to adapt to the era of ‘big data’ in which ‘traditional’ risk factors can be combined with individualised data including genomics and other high-dimensional omics ^4–6^. Coupled with this is evidence that a seemingly disparate range of postoperative complications may be due, in part, to an aberrant host response to the inflammatory insult of surgery ^7–15^. This suggests there could be a common immune-mediated component which is generic rather than organ- or operation-specific; however, whether this is reflected in a shared genetic architecture across different complications remains unknown.

Genome-wide association studies (GWAS) are a powerful approach to study such complex outcome phenotypes. They can provide insight into the underlying mechanisms and biological substrate whilst identifying genetic variants to be used for risk prediction or therapeutic targeting ^16^. GWAS for postoperative complications have examined atrial fibrillation (AF) ^17^, venous thromboembolism (VTE) ^18^, acute kidney injury (AKI) ^19^, and a composite of myocardial infarction (MI), AF, stroke, AKI and delirium ^20^. Most have found no associated variants at a genome-wide significance level but sample size has been a common limitation, as has the use of methods which can be unstable in the context of case-control imbalance ^21^.

To investigate genetic risk factors for postoperative complications and address the methodological limitations of previous studies we conducted a GWAS of postoperative complications after major surgery in the UK Biobank cohort, controlling for case-control imbalance. We focused on AKI, AF, acute MI, stroke and surgical site infection (SSI) as common, possible immune-mediated complications. We then extended our analyses to explore whether these complications share genetic risk factors with one another, and whether the postoperative phenotypes are genetically related to their non-postoperative equivalents.

## Materials and methods

### Study populations

The primary analysis was conducted in UK Biobank, a large-scale biomedical database and research resource containing genetic, lifestyle and health information from half a million UK participants recruited between 2006 and 2010 (http://www.ukbiobank.ac.uk). Linked health data is available for all participants with follow-up to 2022 ^22^. The UK Biobank study was approved by the North-West Multi-centre Research Ethics Committee and all participants provided written informed consent. This research has been conducted under UK Biobank project number 128619.

FinnGen (freeze 9) was used as an external cohort for genetic correlation analyses. FinnGen is a large-scale genomics initiative that has analysed over 500,000 Finnish biobank samples and correlated genetic variation with health data to understand disease mechanisms and predispositions. The project is a collaboration between research organisations and biobanks within Finland and international industry partners ^23^.

### Genotyping

We used UK Biobank genotype data that was previously generated using Affymetrix using two purpose-designed arrays. The genotype data had been quality controlled, phased and ∼96 million genotypes imputed using the Haplotype Reference Consortium and UK10K haplotype resources. FinnGen genotyping used a ThermoFisher Axiom custom chip array with imputation to a reference dataset derived from Finnish whole-genome sequences. Further information on the methods and QC pipeline are available ^23,24^.

### Inclusion and exclusion criteria

We defined eligible participants as those undergoing major, inpatient surgery after their date of enrolment in UK Biobank. Major surgery was defined by OPCS4 codes using the Bupa Schedule of Procedures and Abbott classification ^25,26^. Exclusion criteria for all phenotypes included a previous diagnosis of the outcome of interest and planned day case surgery. For acute MI, cardiothoracic and cardiology procedures were excluded to reduce the risk of the diagnosis preceding the procedure (see Supplementary Methods for more detail on phenotype definition and exclusions).

### Outcome definition

The primary outcome for each GWAS was a first diagnosis of the complication within a 30 day period following surgery. Controls were defined as those undergoing qualifying surgery but not having the outcome within 30 days. Outcome diagnoses were identified using the following International Classification of Diseases, Tenth Revision (ICD-10) codes: atrial fibrillation, I48.9; acute kidney injury, N17; myocardial infarction, I21; stroke, I61, I63 and I64; and surgical site infection, T81.4.

### Genome-wide Association Study (GWAS)

Case-control GWAS adjusted for sex, genetic chip and the first ten genetic principal components was performed for each outcome using REGENIE (v3.2.2) ^21^. Previously established MRC-IEU workflows and methods were used for quality control and to restrict the sample to individuals of European ancestry ^27^. Genome-wide significance was defined as *p* < 5 x 10^-8^ with an additional exploratory threshold of *p* < 1 x 10^-6^. Post-hoc analyses were performed using Functional Mapping and Annotation of Genome-Wide Association Studies (FUMA GWAS ^28^). Independent significant SNPs were defined as those meeting the genome-wide significance threshold and independent of each other at a linkage disequilibrium (LD) threshold of r^2^ < 0.6. Lead SNPs were identified from independent significant SNPs at r^2^ < 0.1. Gene-based analysis was performed using Multi-marker Analysis of GenoMic Annotation (MAGMA ^29^) with a genome-wide significance threshold of p < 0.05/19,840 (2.52 x 10^-6^).

### Sensitivity analyses

The primary GWAS analyses did not include additional clinical covariates as they may be mediators of genetic risk or introduce collider bias given the selection of a surgical patient cohort. The exception to this was postoperative AF, where age was included as an additional covariate due to established age-dependent differences in the contribution of genetic variation ^30^ and subgroup analyses based on cardiothoracic/non-cardiothoracic surgery were performed. Sensitivity analyses were performed with adjustment for age in the other outcomes. Further sensitivity analyses were performed for each outcome with surgical specialty (categorised as cardiothoracic, general, neurosurgery, orthopaedics, vascular or other) included as a covariate to ensure primary results were not driven by specialty-specific enrichment. To evaluate the consistency of genetic effects across clinical contexts, we performed a stratified association analysis of the lead SNPs identified in each primary GWAS to compare effect sizes and directionality. Where there was evidence of potential heterogeneity we undertook a further genotype-by-specialty interaction analysis to test for significant interaction effects between genotype and surgical specialty. Finally, sensitivity GWAS were undertaken with a postoperative washout period of 180 days for controls to improve phenotypic separation.

### Genetic correlation analyses between postoperative complications

The GWAS results were used to analyse genetic correlation across phenotypes. Global genetic correlation was assessed through cross-trait LD Score Regression ^31,32^ and Local Analysis of [co]Variant Association (LAVA) was used to analyse bivariate local genetic correlation in 2,495 predefined regions ^33^.

### Genetic correlation analysis with non-postoperative phenotypes

GWAS summary statistics were obtained from the FinnGen study cohort (Freeze 9, https://r9.finngen.fi/) for non-postoperative equivalents of the outcome phenotypes, i.e. undifferentiated AF, AKI, MI and stroke, as well as chronic kidney disease (CKD). SSI was excluded as there is no suitable non-postoperative phenotype. The results of the primary GWAS analyses were used with this external dataset to analyse global and local genetic correlation between the postoperative and non-postoperative paired phenotypes as above.

### Polygenic risk score analysis

Polygenic risk scores (PRS) were identified in the Polygenic Score Catalog (https://www.pgscatalog.org) for non-postoperative AF, AKI/CKD, MI and stroke. No PRS were available for SSI. The *pgsc_calc* workflow ^34^ was used to download, variant match and calculate individual polygenic scores for the UK Biobank cohort. Scores were included if at least 75% of variants present in the score were also present in the UK Biobank genomic data. To provide a consensus estimate of genetic liability whilst mitigating selection bias and reducing the risk of overfitting we employed an ensemble approach. Individual scores which passed matching were centred and scaled to ensure comparability across different scoring methodologies. For each outcome, these scaled scores were then summed and averaged to create a single ensemble risk score. The resulting average score was then converted into risk quintiles (1=lowest risk, 5=highest) to improve interpretability ^35^. Odds ratios for the association of increasing quintile of PRS for the non-postoperative phenotype with the postoperative equivalent were estimated by logistic regression after adjustment for age and sex using the *glm* function in R (version 4.4.0). To assess the sensitivity of our findings to score selection we performed two additional analyses for each phenotype using a) the single score with the highest reported performance (AUROC or C-statistic) in the original publication and b) the score developed in the largest discovery sample size.

## Results

### Participant characteristics

A total of 8,472 complications (AF, AKI, MI, SSI, or stroke) occurred in 140,563 eligible individuals. Case-control numbers for each outcome phenotype after excluding previous diagnoses (and cardiothoracic/cardiology procedures for AMI) are shown in Table 1. Most individuals (7,711) had a single complication with 701 and 60 individuals having two and three complications respectively. The most common combinations of complications were AF and AKI (228 individuals) and AKI and SSI (205 individuals).

**Table 1.** Numbers of cases and controls for each outcome phenotype in UK Biobank.

| **Outcome** | **Cases** | **Controls** |
| --- | --- | --- |
| Acute kidney injury | 3,002 | 134,941 |
| Atrial fibrillation | 2,739 | 135,878 |
| Surgical site infection | 2,701 | 136,120 |
| Stroke | 569 | 137,842 |
| Acute myocardial infarction | 282 | 123,339 |

Individuals who developed postoperative complications tended to be older, were more likely to be male, current or previous smokers and had higher rates of comorbidity (all p < 0.001) (Table 2). Compared to controls, cases were more likely to be emergency admissions (2,899 (34%) vs 27,690 (21%), p < 0.001) and to have undergone cardiothoracic surgical procedures (1,158 (14%) vs 4,211 (3.2%), p < 0.001). Additional details and tables of participant characteristics for each outcome phenotype can be found in supplementary tables 1-6.

**Table 2.** Summary of participant characteristics for cases and controls. For individuals experiencing multiple complications, details of the first complication were used. For controls, details of the first eligible surgical procedure were used.

|  | **Cases**<br>**n = 8,472** | **Controls**<br>**n = 132,091** | **p-value\*** |
| --- | --- | --- | --- |
| **Age (yr), mean (SD)** | 68 (6) | 64 (8) | $<2.2 \times 10^{-16}$ |
| **Female sex, number (%)** | 3,524 (42) | 74,283 (56) | $4.7 \times 10^{-152}$ |
| **Ethnicity, number (%)** |  |  | 0.79 |
| **Asian or Asian British** | 146 (1.7) | 2,166 (1.6) |  |
| **Black or Black British** | 94 (1.1) | 1,565 (1.2) |  |
| **Chinese** | 12 (0.1) | 228 (0.2) |  |
| **Mixed** | 36 (0.4) | 644 (0.5) |  |
| **Other ethnic group** | 54 (0.6) | 935 (0.7) |  |
| **White** | 8,090 (96) | 125,951 (96) |  |
| **Missing** | 40 (0.5) | 602 (0.5) |  |
| **Smoking status, number (%)** | | | $3.8 \times 10^{-37}$ |
| **Current** | 1,138 (13) | 14,151 (11) |  |
| **Never** | 3,697 (44) | 66,825 (51) |  |
| **Previous** | 3,570 (42) | 50,389 (38) |  |
| **Prefer not to answer or missing** | 67 (0.8) | 726 (0.5) |  |
| **Charlson Comorbidity Index, median IQR)** | 1.00 (0.00-2.00) | 0.00 (0.00-1.00) | $<2.2 \times 10^{-16}$ |
| **Admission method, number (%)** | | | $8.1 \times 10^{-224}$ |
| **Elective** | 5,242 (62) | 101,732 (77) |  |
| **Emergency** | 2,899 (34) | 27,690 (21) |  |
| **Other** | 331 (3.9) | 2,669 (2) |  |
| **Surgical specialty, number (%)** | | | $<2.2 \times 10^{-16}$ |
| **Cardiothoracic surgery** | 1,158 (14) | 4,211 (3.2) |  |
| **General surgery** | 2,229 (26) | 24,604 (19) |  |
| **Neurosurgery** | 371 (4.4) | 4,902 (3.7) |  |
| **Other** | 1,865 (22) | 34,937 (26) |  |
| **Trauma and orthopaedics** | 1,986 (23) | 51,927 (39) |  |
| **Urology** | 624 (7.4) | 10,080 (7.6) |  |
| **Vascular** | 239 (2.8) | 1,430 (1.1) |  |

Numbers are number (proportion), mean (SD) or median (IQR). Additional details can be found in Supplementary table 1. \*p-values reported as $<2.2 \times 10^{-16}$ were below estimable limit of the relevant R function.

### Genome-wide association study: postoperative atrial fibrillation

A total of 171 genome-wide significant variants were identified for postoperative AF, with nine independent significant SNPs across three genomic risk loci on chromosomes 1, 4 and 16 identified by FUMA GWAS (Figures 1–2; Table 3). The lead SNPs were rs36088503, 4:111697815_TG_T (GRCh37.p13) and rs145672192 respectively. The nearest genes were *KCNN3*, which reached genome-wide significance in a MAGMA gene-level analysis (p = 2.4 x 10^-7^, Figure 3), *PITX2, RP11-777N19.1* and *TRAF7:CASKIN1*. A further 38 genes were mapped to these regions (supplementary table 7).

**Figure 1.**
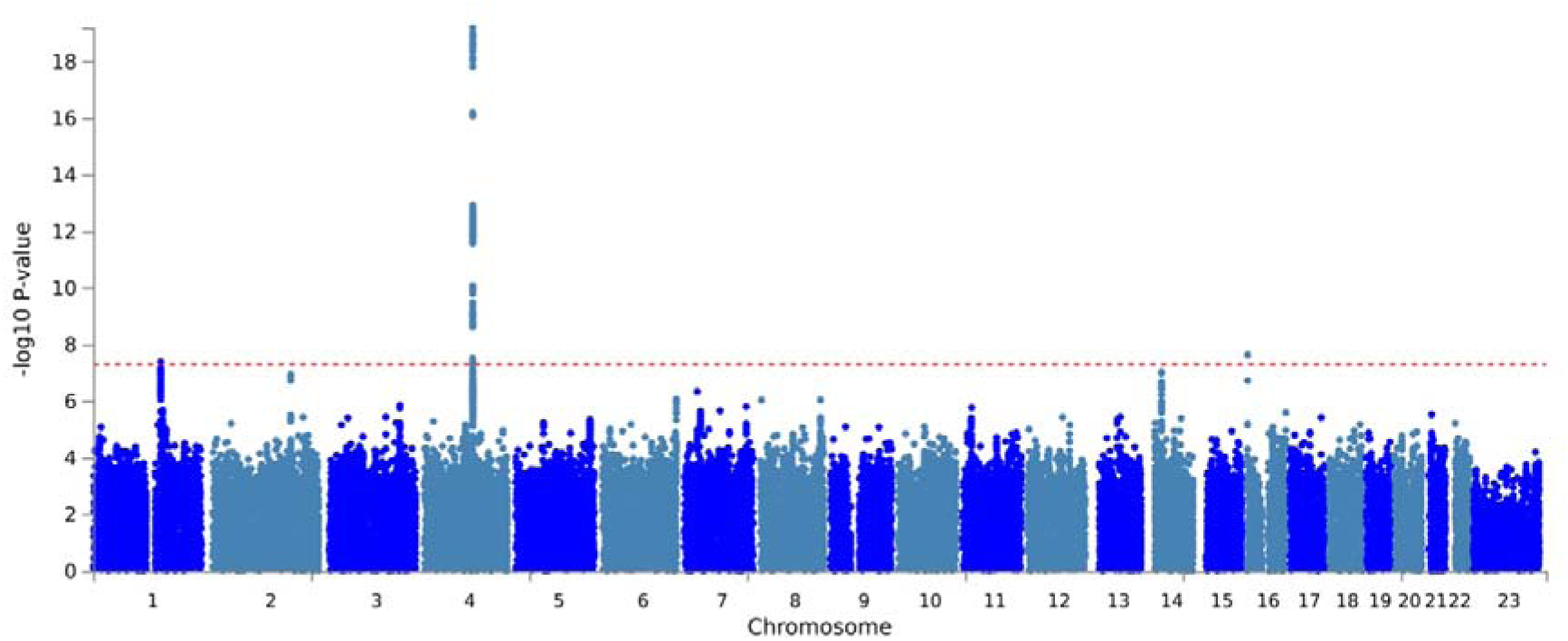
Manhattan plot of genome-wide association study results for postoperative atrial fibrillation. The red line represents a genome-wide significant p value of 5 x 10^-8^.

**Figure 2.**
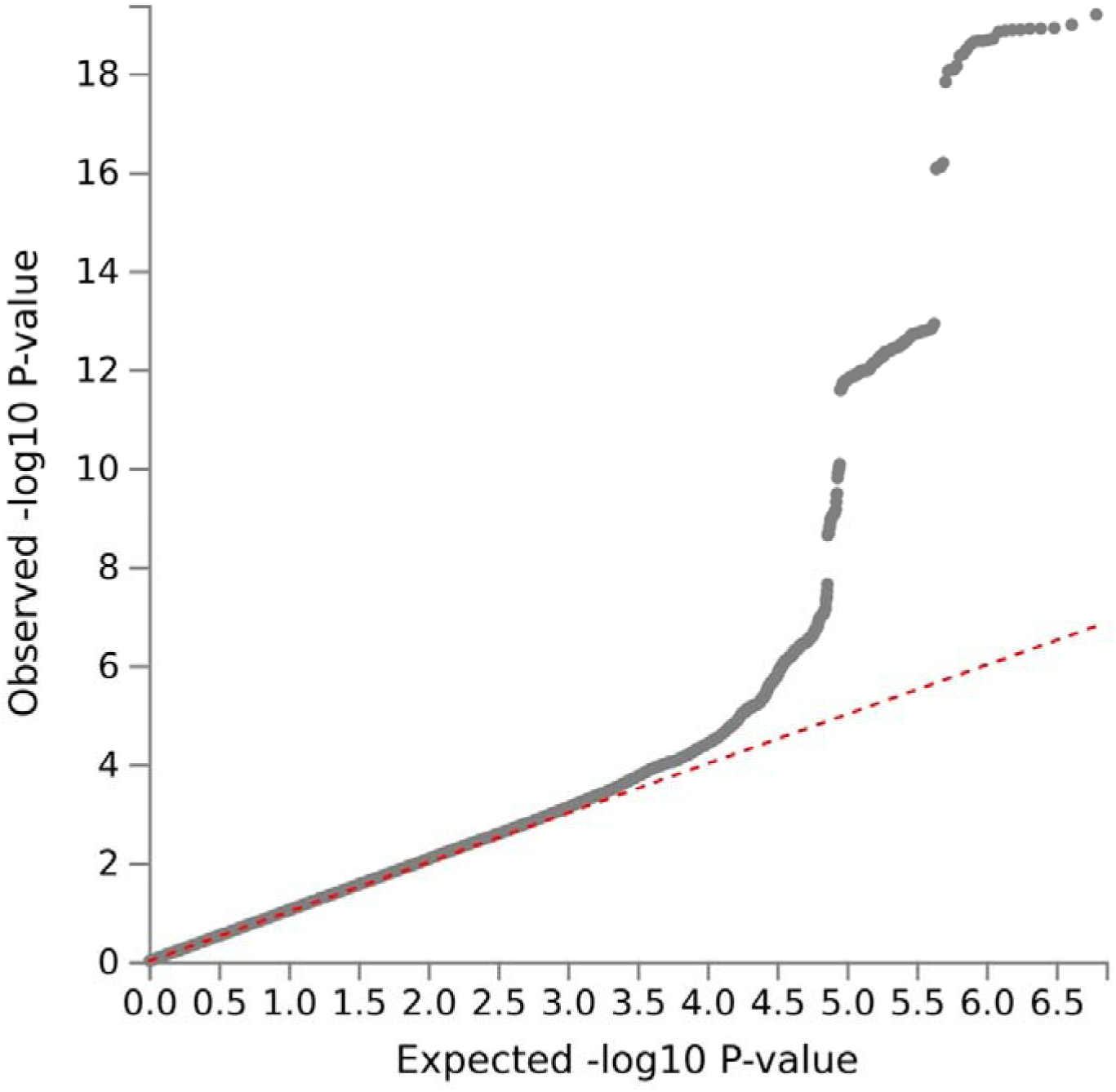
Q-Q plot for genome-wide association study of postoperative atrial fibrillation. Genomic inflation factor (λ) 1.04.

**Figure 3.**
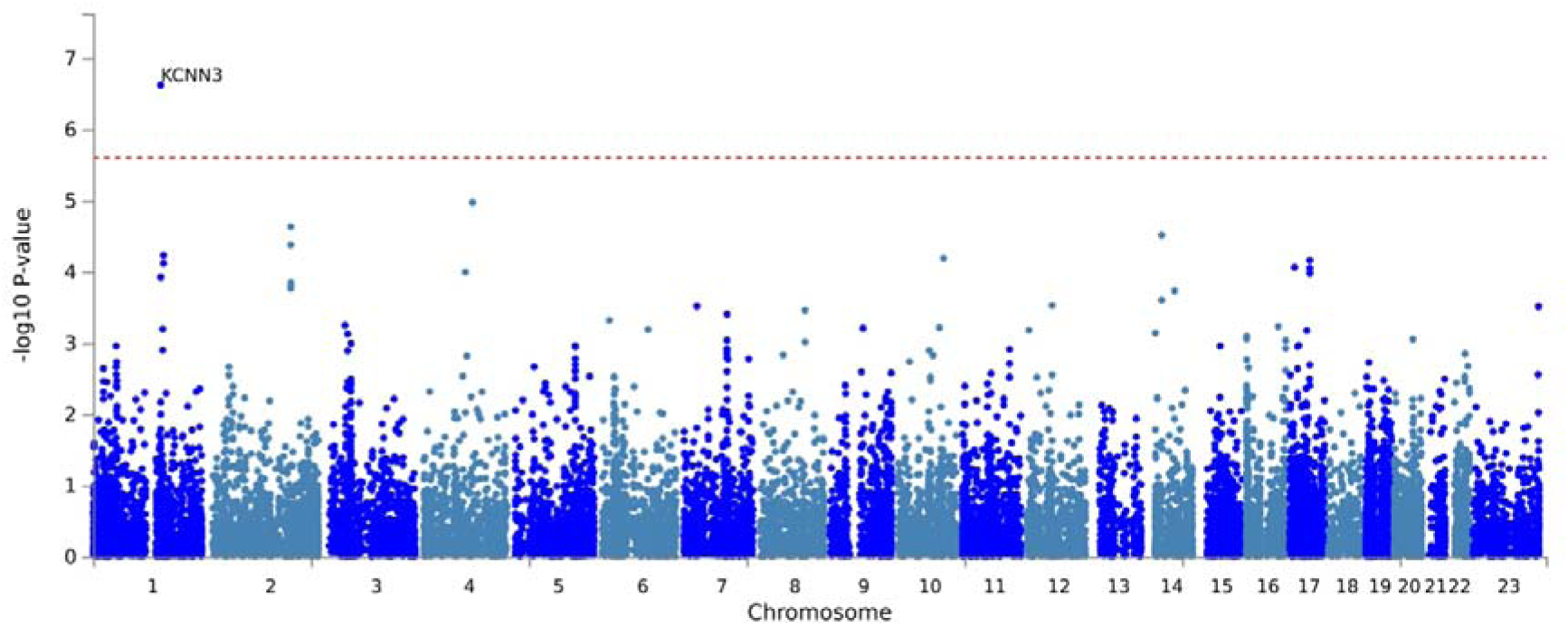
Manhattan plot of gene-based testing for postoperative atrial fibrillation

**Table 3.**
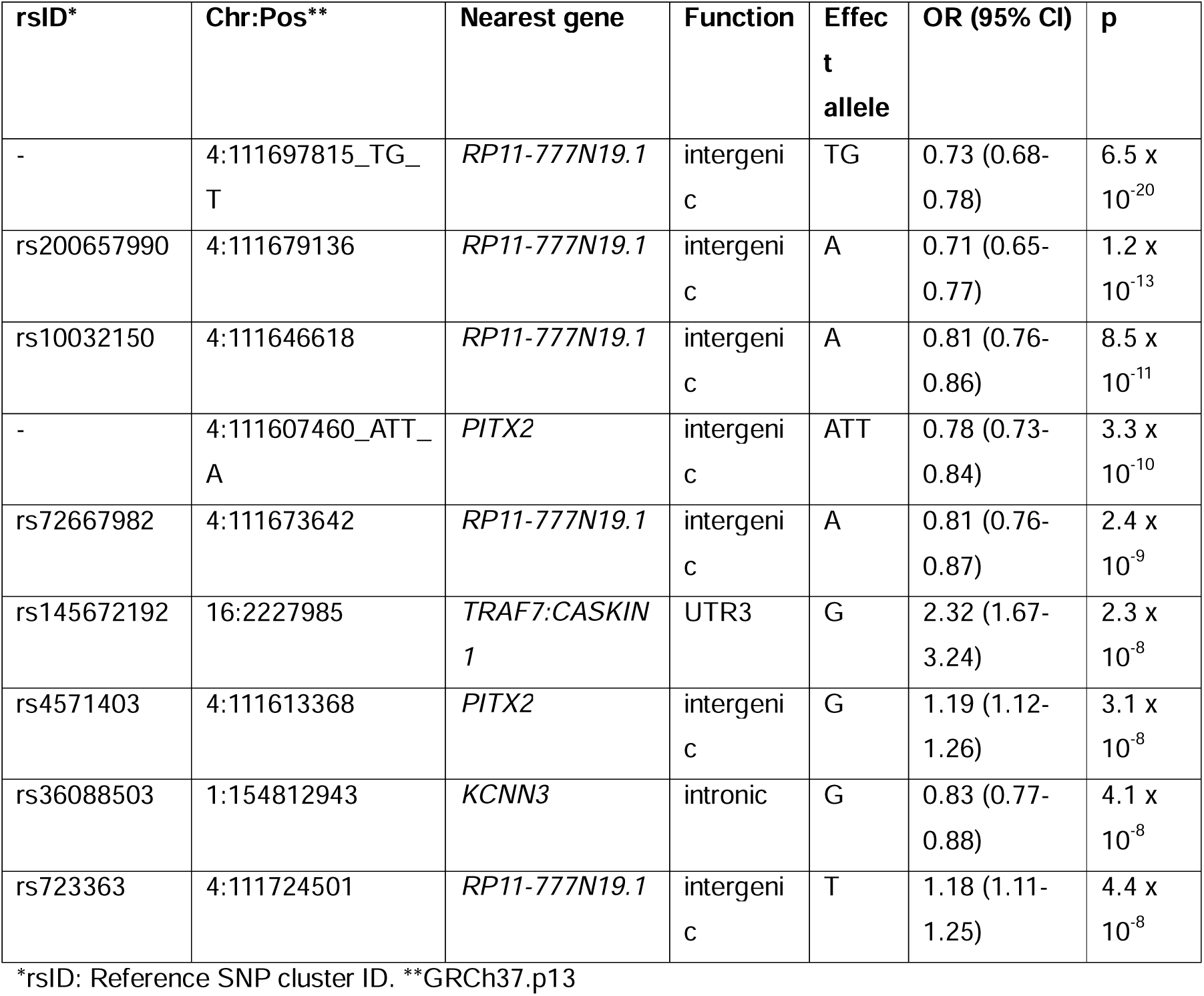
Independent significant SNPs identified in case-control GWAS of postoperative atrial fibrillation.

| rsID* | Chr:Pos** | Nearest gene | Function | Effect allele | OR (95% CI) | p |
| --- | --- | --- | --- | --- | --- | --- |
| - | 4:111697815_TG_T | *RP11-777N19.1* | intergenic | TG | 0.73 (0.68-0.78) | 6.5 x 10<sup>-20</sup> |
| rs200657990 | 4:111679136 | *RP11-777N19.1* | intergenic | A | 0.71 (0.65-0.77) | 1.2 x 10<sup>-13</sup> |
| rs10032150 | 4:111646618 | *RP11-777N19.1* | intergenic | A | 0.81 (0.76-0.86) | 8.5 x 10<sup>-11</sup> |
| - | 4:111607460_ATT_A | *PITX2* | intergenic | ATT | 0.78 (0.73-0.84) | 3.3 x 10<sup>-10</sup> |
| rs72667982 | 4:111673642 | *RP11-777N19.1* | intergenic | A | 0.81 (0.76-0.87) | 2.4 x 10<sup>-9</sup> |
| rs145672192 | 16:2227985 | *TRAF7:CASKIN1* | UTR3 | G | 2.32 (1.67-3.24) | 2.3 x 10<sup>-8</sup> |
| rs4571403 | 4:111613368 | *PITX2* | intergenic | G | 1.19 (1.12-1.26) | 3.1 x 10<sup>-8</sup> |
| rs36088503 | 1:154812943 | *KCNN3* | intronic | G | 0.83 (0.77-0.88) | 4.1 x 10<sup>-8</sup> |
| rs723363 | 4:111724501 | *RP11-777N19.1* | intergenic | T | 1.18 (1.11-1.25) | 4.4 x 10<sup>-8</sup> |

\*rsID: Reference SNP cluster ID. \*\*GRCh37.p13

The primary findings were robust to a control washout period of 180 days with eight independent significant SNPs returned across the same risk loci on chromosomes 4 and 16 (supplementary table 8). The lead SNP on chromosome 1 in the primary analysis, rs36088503, showed consistent effects though the p-value was marginally below the genome-wide significance threshold (OR (95% CI) sensitivity GWAS 0.83 (077–0.88) vs primary GWAS 0.83 (0.77–0.88), p = 5.3 x 10^-8^). In the sensitivity analysis adjusting for surgical specialty, eight independent significant SNPs were identified across the primary risk loci on chromosomes 1 and 4 (supplementary table 9). While the lead SNP on chromosome 16 (rs145672192) showed a stable effect size (adjusted OR (95% CI) 2.3 (1.2–3.2) vs primary OR 2.3 (1.7–3.2)), it no longer met the strict genome-wide significance threshold (p = 1.2 x 10^-7^).

Subgroup analyses were performed in cardiothoracic and non-cardiothoracic surgery cohorts. No genome-wide significant SNPs or genes were identified for the cardiac surgery cohort (962 cases, 13,504 controls). For the non-cardiac cohort (1777 cases, 122,374 controls), six independent significant SNPs were identified in risk loci on chromosomes 2 and 4 from a total of 162 genome-wide significant variants (supplementary table 10). The results for rs145672192 (chromosome 16) mirrored the specialty-adjusted GWAS, showing consistent effects (OR (95% CI) 2.8 (1.9–4.4)) but not reaching genome-wide significance (p = 1.4 x 10^-7^).

Stratified association analysis of the primary GWAS lead SNPs confirmed that the effect of the rs145672192 was robust in orthopaedic, general and other surgical specialties, but was absent in cardiac and vascular cohorts (supplementary figure 9). The genotype-by-specialty interaction analysis confirmed a significant interaction effect between rs145672192 and cardiac vs non-cardiac surgical status (p = 0.007) suggesting the contribution of rs145672192 is diminished or masked in the context of cardiac surgery (supplementary table 11).

### Genome-wide association study: other complications

No genome-wide significant SNPs were found for AKI, AMI, stroke or surgical site infection (supplementary figures 1–8) in the primary or sensitivity analyses. At an exploratory threshold of p < 1 x 10^-6^, three independent variants were identified for both AKI and AMI, and eight for SSI (supplementary table 12). Stratified analysis of these exploratory SNPs demonstrated consistent directionality across surgical specialties, though the stability of the effect sizes varied with subgroup power (supplementary figures 10-12).

### Genetic correlation between postoperative complications in UK Biobank

No significant global genetic correlation was found between the different postoperative phenotypes in UK Biobank on cross-trait LD Score Regression analysis (supplementary table 13). Local genetic correlation between complications was therefore not assessed given the low SNP-based heritability z-scores and effective sample sizes ^31^.

### Genetic correlation with between postoperative (UK Biobank) and non-postoperative (FinnGen) phenotypes

A strong global genetic correlation was observed between postoperative AF in UK Biobank and non-postoperative AF in the FinnGen cohort (*r_g_* = 0.69, p = 0.001). Significant local correlation was confirmed in two regions near to the *KCNN3* (Chr 1) and *PITX2* (Chr 4) genes identified in the primary GWAS (Table 4). There was some suggestion of shared genetic liability between postoperative AKI and chronic renal failure, but the evidence for global genetic correlation was limited (*r_g_* = 0.42, p = 0.05) and there was little evidence of local genetic correlation. The results for the other phenotypes did not support the presence of global genetic correlation between postoperative and non-postoperative settings (supplementary table 14) and were not analysed further.

**Table 4.** Regions of significant local genetic correlation between postoperative atrial fibrillation (in UK Biobank) and non-postoperative atrial fibrillation (in FinnGen)

| Chr | Start | Stop | Rho | R<sup>2</sup> | p |
| --- | --- | --- | --- | --- | --- |
| 1 | 154685546 | 156813845 | 0.54 (0.36-0.74) | 0.29 (0.13-0.55) | $7.3 \times 10^{-8}$ |
| 4 | 109978983 | 111733579 | 0.58 (0.44-0.73) | 0.34 (0.20-0.45) | $2.1 \times 10^{-13}$ |

*Rho is the estimate of the local genetic correlation between traits (moderate to strong positive correlation: genetic variants in this region may affect both traits in a similar way). R<sup>2</sup> is the proportion of variance explained by the local genetic correlation, or how much of the trait variability can be account for by this region. Proportion of covariance between the traits that is explained by variation in this range.*

### Association of polygenic risk scores for related phenotypes (PGS Catalog) with postoperative complications (UK Biobank)

Ensemble polygenic risk scores were calculated for non-postoperative AF (20 individual PRS), AMI (6 PRS) and stroke (25 PRS). No PRS were available for AKI but 20 scores were identified for chronic kidney disease (supplementary table 15). After adjusting for age and sex, increasing PRS quintile was associated with increased odds of postoperative complications for all phenotypes with the most pronounced effects seen for AF and AMI (Figure 4). The AUC (95% CI) for the full models ranged from 0.63 (0.61–0.66) for stroke to 0.83 (0.82–0.84) for AF (supplementary table 16). Sensitivity analyses comparing the ensemble PRS approach to single scores selected by best reported performance (supplementary table 17) or largest discovery sample size (supplementary table 18) demonstrated the observed associations were robust. While intermediate quintiles showed some attenuation in single-score models, the association for individuals in the highest risk quintile remained significant across all outcomes. In a subgroup analysis for AF stratified by cardiothoracic/non-cardiothoracic surgery, stronger effects were seen in the non-cardiothoracic cohort: PRS quintile 5, OR (95% CI) for non-cardiothoracic surgery, 6.5 (5.4–7.8), and for cardiothoracic surgery 2.6 (2.1–3.3) (supplementary table 19).

**Figure 4.**
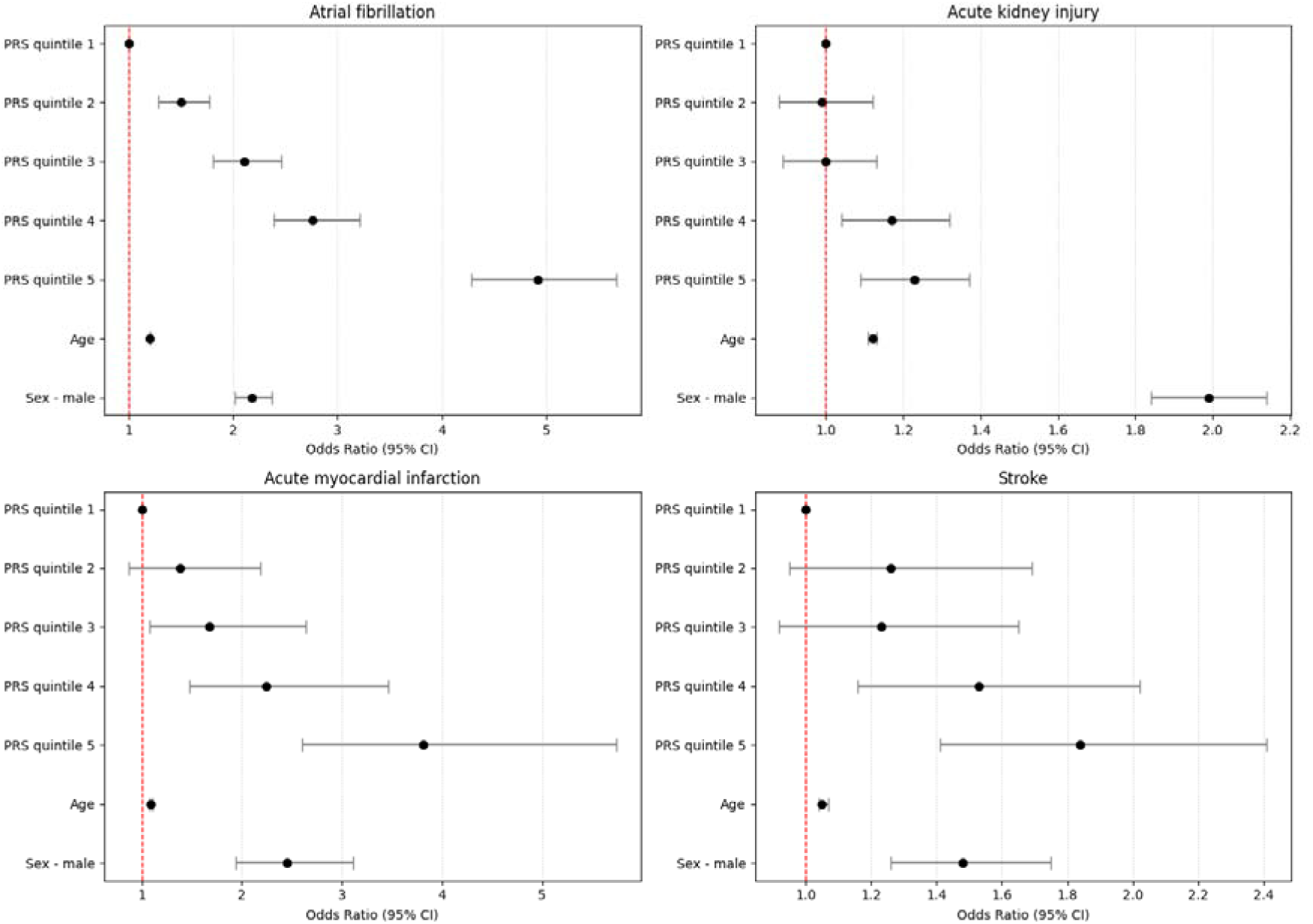
Association between postoperative complications and increasing polygenic risk score quintile for non-postoperative equivalent phenotype. Red line is odds ratio of 1. Error bars are 95% confidence interval.

## Discussion

In this study we used a UK-based cohort with linked healthcare data to explore the genetic basis of common postoperative complications. We identified three genomic loci associated with postoperative atrial fibrillation (AF) and evidence of shared genetic risk with non-postoperative AF. No genome-wide significant risk loci were identified for the other outcomes studied, and there was no evidence of shared genetic risk between postoperative complications.

Our study is better powered than previous GWAS of postoperative AF (number of cases/controls: 257/620 and 242/640) which failed to find any genome-wide significant results ^36,37^. A comparable previous GWAS of postoperative AF using the UK Biobank cohort reported two variants in proximity to the *PITX2* gene on chromosome 4 ^17^. However, our analysis builds on these findings by restricting eligibility to major surgical procedures, defining the outcome phenotype more strictly, and employing a method that is robust in the context of case-control imbalance ^21^. We identified independent significant SNPs in a high degree of linkage disequilibrium (LD) with those variants, adding to the evidence implicating the 4q25 locus and *PITX2* gene, established genetic predictors of AF more broadly, in postoperative AF ^38–40^. We also found additional risk loci on chromosomes 1 and 16. The 1q21 region has previously been associated with non-postoperative AF in GWAS ^40^ and post-cardiac surgery AF in a targeted genotyping analysis ^39^. We found this region, including the *KCNN3* gene, to be a risk locus for postoperative AF with additional evidence of genetic correlation with non-postoperative AF.

The third risk locus identified, 16p13 (lead SNP rs145672192), has not previously been reported in association with postoperative AF. It is distinct from the established AF risk locus 16q22 (*ZFHX3*) and is not in LD with it. Whilst the proximity of *TRAF7:CASKIN1*, coregulated genes with roles in inflammatory signalling (*TRAF7* ^41^) and cardiac excitability (*CASKIN1* ^42^), makes this a biologically plausible candidate locus, the finding should be considered hypothesis-generating given the current lack of functional validation or external replication. The biological relevance of this locus is supported by the specialty-stratified and genotype-by-specialty interaction analyses. The observed effect of rs145672192 was robust in non-cardiac procedures but attenuated in the cardiac surgery cohort, similar to the additional lead SNP on chromosome 2 (rs12052983) identified in the non-cardiac subgroup GWAS but not in the primary analysis. These findings, alongside the results of our genetic and PRS-based analyses, support the distinction between AF after cardiac surgery and postoperative AF in other contexts, and suggest that the genetic influence on postoperative AF is particularly marked in non-cardiac surgery. The unique set of pathophysiological contributory mechanisms in the cardiac surgery setting, for example direct atrial manipulation and cardiopulmonary bypass, may overwhelm the more subtle effect of genetic predisposition and make a reduced genetic influence a plausible finding ^46^.

We found no genome-wide significant variants associated with postoperative AKI, MI, stroke or SSI and no global genetic correlation between the different postoperative phenotypes, suggesting that a singular shared genetic architecture is unlikely. Whilst the study was better powered than a previous GWAS including AKI, MI and stroke ^37^ we were underpowered to detect small effects (supplementary table 20). The post-hoc power for AKI and SSI were similar to that of AF and so it may be that these outcomes are driven more by environmental or direct perioperative factors than underlying genetic susceptibility. This is supported by SNP-based heritability estimates of 22.1% for AF ^43^ versus <0.1% and 4.5% for AKI after cardiac and non-cardiac surgery respectively ^44,45^.

The relationship between postoperative and non-postoperative paired phenotypes was explored by genetic correlation and polygenic risk score analyses. We found suggestive evidence of global genetic correlation between AKI and CKD as well as increased risk of postoperative AKI with increasing PRS for CKD. Whilst the negligible SNP-based heritability for MI and stroke prevented an assessment of genetic correlation between the postoperative and non-postoperative equivalents, the PRS analyses demonstrated that increasing PRS for non-postoperative MI and stroke were associated with increased risk of the relevant postoperative complication. Taken together, these findings may indicate a ‘two-hit’ paradigm in which the inflammatory insult of surgery precipitates the outcome in individuals with underlying genetic vulnerability for related chronic pathology via mechanisms which appear predominantly trait-specific rather than generalised across outcomes.

There may therefore be a role for existing PRS in risk-stratification and preventive perioperative care. In the case of postoperative AF, a review of clinical practice across UK cardiac surgery centres found most did not risk-stratify patients or have a care package in place to prevent postoperative AF ^47^. The use of PRS for non-postoperative AF could therefore be a useful adjunct in the pre-operative phase, highlighting those at highest risk who may benefit from a preventive package of care. There are also implications for the longer-term follow-up of acute postoperative complications. Given the overlap of polygenic risk between these postoperative phenotypes and their non-postoperative equivalents, there may be a proportion of individuals in whom the acute postoperative event is the initial presentation of undiagnosed pathology.

This study does have limitations. Some factors which would be of clinical relevance are not collected in UK Biobank or the linked NHS healthcare data, for example ASA physical status grade or type of anaesthesia. Additionally, the UK Biobank cohort is not representative of the wider population, particularly due to differences in socioeconomic status and comorbid status ^48^. The postoperative complications studied were defined by clinical coding data and were not prospectively screened for. This may have resulted in some cases being missed; however, this would dilute any observed effect and so suggests our findings are robust to this misclassification. Whilst efforts were taken to minimise the risk of the outcomes preceding surgery it is possible that some cases remain. The limited sample size, particularly for some outcomes, restricted the power of our primary analysis and the application of some genetic correlation methods. Finally, the external cohort used does not define postoperative phenotypes and so direct replication and validation of the 16p13 locus was not possible.

In conclusion, we found multiple genomic risk loci associated with postoperative atrial fibrillation in a major surgery cohort, including a putative novel locus on chromosome 16 which warrants further investigation and replication. Our findings suggest that for certain complications, most notably AF, there is shared genetic aetiology between the postoperative phenotype and its non-postoperative equivalent. Whilst we did not find evidence of a common genetic mechanism spanning distinct postoperative complications, the association between existing polygenic risk scores and several postoperative outcomes suggests that underlying chronic disease liability contributes to perioperative risk of acute presentations.

## Data Availability

Summary statistics will be publicly available on publication. Source data are available from UK Biobank (http://www.ukbiobank.ac.uk) and Finngen (https://r9.finngen.fi) through their access procedures.
Author-generated code to support the manuscript is available on GitHub (https://doi.org/10.5281/zenodo.17901910)

## Acknowledgements

This research has been conducted using the UK Biobank Resource under Application Number 128619. This includes linked data provided by patients and collected by the NHS as part of their care and support and data assets made available by National Safe Haven as part of the Data and Connectivity National Core Study, led by Health Data Research UK in partnership with the Office for National Statistics and funded by UK Research and Innovation (grant ref MC_PC_20058).

Quality Control filtering of the UK Biobank data used the process described by R.Mitchell, G.Hemani, T.Dudding, L.Corbin, S.Harrison, L.Paternoster (https://data.bris.ac.uk/data/dataset/1ovaau5sxunp2cv8rcy88688v).

This work was carried out using the computational and data storage facilities of the Advanced Computing Research Centre, University of Bristol (http://www.bristol.ac.uk/acrc/).

For the purpose of open access, the author(s) has applied a Creative Commons Attribution (CC BY) licence to any Author Accepted Manuscript version arising from this submission.

## Funding Statement

RAA is funded by a Wellcome Trust GW4-CAT PhD Programme for Health Professionals PhD Fellowship [316275/Z/24/Z]. RAA, PY, GMK and TRG are supported by the Medical Research Council Integrative Epidemiology Unit at the University of Bristol (RA, TG: MC_UU_00032/3; PY: MC_UU_00032/4; GMK: MC_UU_0032/6). GMK acknowledges additional funding from the Wellcome Trust (grant numbers: 201486/Z/16/Z and 201486/B/16/Z), the Medical Research Council (grant numbers: MR/W014416/1; MR/S037675/1; MR/Z50354X/1; and MR/Z503745/1. PY, TRG, and GMK are also supported by the UK National Institute for Health and Care Research (NIHR) Bristol Biomedical Research Centre (grant number: NIHR 203315). The views expressed are those of the authors and not necessarily those of the UK NIHR or the Department of Health and Social Care.

## Conflicts of Interest

TRG receives funding from GlaxoSmithKline, Biogen and Roche for unrelated research. The other authors declare that they have no conflict of interest.

## Notes

### Author Declarations

The primary analysis was conducted in UK Biobank. The UK Biobank study was approved by the North-West Multi-centre Research Ethics Committee and all participants provided written informed consent. This research has been conducted under UK Biobank project number 128619.

### Summary of Updates

Revision after initial journal submission and peer-review. Several new sensitivity analyses with changes throughout.

