## Supplementary material for "Genetic risk factors for postoperative complications after major surgery and shared genetic aetiology with non-postoperative phenotypes"

**Supplementary methods**

UK Biobank (UKB) contains linked Hospital Episode Statistics (HES) inpatient data for participants. The diagnosis table contains all diagnoses (ICD-10 code) associated with each admission episode. Individuals with a previous diagnosis of the outcome phenotype of interest were excluded to reduce potential bias through confounding and reverse causation. It also ensured all events occurred after baseline variables were measured at UKB enrolment and contributed to a more homogenous case-control cohort.

The operation table contains details of all OPCS4 procedure codes for a patient during a given admission. Inpatient surgery was defined according to patient class (excluding day case). Major surgery was defined by using a combination of two grading systems from the literature (Bupa schedule of procedures(1) and Abbott(2)). Eligible procedures were either Bupa ‘major’ or ‘complex’ category or, if not classified in Bupa, in the ‘restrictive’ category as defined by Abbott. Only procedures performed after enrolment in UKB were eligible. All surgical specialties were included except for the acute myocardial infarction cohort, where cardiothoracic and cardiology procedures were excluded.

In cases of multiple procedures on the same date, the procedure coded as level 1 was treated as the index procedure. If there were multiple or no level 1 procedures, the procedure with the highest grade was used (complex > major > restrictive). If there were multiple procedures with the highest grade, minimum array index was used as a tiebreak. Count variables were added to account for a) multiple surgical procedures within the same hospital admission and b) cumulative procedures since UKB enrolment.

A limitation of HES coding is that procedures have a specific date of operation, whereas diagnoses are linked to an entire inpatient episode (without a specific date of diagnosis). To limit the risk of a diagnosis preceding an operative procedure where both occurred in the same inpatient episode, the following decision tree was used:

Complication diagnosis in later episode than operation date?

Yes: include

No

Operation and diagnosis in same episode. Time from episode start to operation <14 days?

Yes

Operation date in first half of episode?

Yes: include

No: exclude

No: exclude

**Supplementary tables**

1. **Additional participant characteristics**
2. **Participant characteristics – atrial fibrillation**
3. **Participant characteristics – acute kidney injury**
4. **Participant characteristics – acute myocardial infarction**
5. **Participant characteristics – stroke**
6. **Participant characteristics – surgical site infection**
7. **Mapped genes from GWAS of postoperative atrial fibrillation**
8. **Clean controls AF**
9. **Specialty-adjusted AF**
10. **Independent significant SNPs from GWAS of postoperative atrial fibrillation after non-cardiothoracic surgery**
11. **Genotype-by-specialty (G x E) interaction results for postoperative atrial fibrillation primary GWAS lead SNPs**
12. **Independent SNPs meeting an exploratory p-value threshold of 1 x 10^-6^ for postoperative acute kidney injury, myocardial infarction and surgical site infection**
13. **Cross-trait genetic correlation results for all postoperative phenotypes**
14. **Cross-trait genetic correlation results between postoperative (UK Biobank) and ambulatory (FinnGen) phenotypes**
15. **Polygenic risk scores passing matching**
16. **Odds ratios for postoperative complication by polygenic risk score quintile**
17. **Odds ratios for postoperative complication by polygenic risk score quintile. Single score for each outcome based on best reported development performance.**
18. **Odds ratios for postoperative complication by polygenic risk score quintile. Single score for each outcome based on largest development sample size.**
19. **Association between polygenic risk score for atrial fibrillation and postoperative atrial fibrillation in cardiothoracic and non-cardiothoracic surgery subgroups**
20. **Post hoc power calculation for primary GWAS**

**Supplementary table 1** Additional participant characteristics – all outcomes

| **Characteristic** | **Case N = 8,472*^1^*** | **Control N = 132,091*^1^*** | **p-value*^2^*** |
| --- | --- | --- | --- |
| Age | 68 (8) | 64 (8) | <2.2E-16 |
| Sex |  |  | 4.69E-152 |
| Female | 3,524 (42) | 74,283 (56) |  |
| Male | 4,948 (58) | 57,808 (44) |  |
| BMI (kg/m2) | 28.5 (25.5, 32.0) | 27.5 (24.7, 30.8) | 1.38E-68 |
| Missing | 49 (0.6%) | 678 (0.5%) |  |
| Ethnicity |  |  | 0.79 |
| Asian or Asian British | 146 (1.7) | 2,166 (1.6) |  |
| Black or Black British | 94 (1.1) | 1,565 (1.2) |  |
| Chinese | 12 (0.1) | 228 (0.2) |  |
| Mixed | 36 (0.4) | 644 (0.5) |  |
| Other ethnic group | 54 (0.6) | 935 (0.7) |  |
| White | 8,090 (96) | 125,951 (96) |  |
| Missing | 40 (0.5%) | 602 (0.5%) |  |
| Townsend deprivation index | -1.9 (-3.6, 0.9) | -2.1 (-3.6, 0.6) | 8.44E-06 |
| Missing | 8 (<0.1%) | 151 (0.1%) |  |
| Alcohol intake |  |  | 3.36E-07 |
| Daily or almost daily | 1,848 (22) | 26,417 (20) |  |
| Never | 821 (9.7) | 11,727 (8.9) |  |
| Once or twice a week | 2,138 (25) | 33,476 (25) |  |
| One to three times a month | 907 (11) | 14,803 (11) |  |
| Prefer not to answer | 16 (0.2) | 154 (0.1) |  |
| Special occasions only | 1,083 (13) | 16,887 (13) |  |
| Three or four times a week | 1,649 (19) | 28,494 (22) |  |
| Missing | 10 (0.1%) | 133 (0.1%) |  |
| Smoking status |  |  | 3.75E-37 |
| Current | 1,138 (13) | 14,151 (11) |  |
| Never | 3,697 (44) | 66,825 (51) |  |
| Prefer not to answer | 57 (0.7) | 594 (0.5) |  |
| Previous | 3,570 (42) | 50,389 (38) |  |
| Missing | 10 (0.1%) | 132 (<0.1%) |  |
| Physical activity (days/week) | 4.00 (2.00, 6.00) | 4.00 (2.00, 6.00) | 0.41 |
| Missing | 579 (6.8%) | 8,330 (6.3%) |  |
| Myocardial infarction | 923 (11) | 5,852 (5.0) | 2.10E-131 |
| Missing | 342 (4.0%) | 15,292 (12%) |  |
| Congestive cardiac failure | 692 (8.5) | 2,837 (2.4) | 2.55E-224 |
| Missing | 342 (4.0%) | 15,292 (12%) |  |
| Cerebrovascular disease | 638 (7.8) | 4,289 (3.7) | 8.17E-78 |
| Missing | 342 (4.0%) | 15,292 (12%) |  |
| Dementia | 78 (1.0) | 434 (0.4) | 2.16E-15 |
| Missing | 342 (4.0%) | 15,292 (12%) |  |
| Chronic pulmonary disease | 1,660 (20) | 14,471 (12) | 1.34E-96 |
| Missing | 342 (4.0%) | 15,292 (12%) |  |
| Diabetes | 1,200 (15) | 8,723 (7.5) | 5.35E-122 |
| Missing | 342 (4.0%) | 15,292 (12%) |  |
| Diabetes with complications | 233 (2.9) | 1,135 (1.0) | 2.52E-56 |
| Missing | 342 (4.0%) | 15,292 (12%) |  |
| Renal disease | 736 (9.1) | 3,292 (2.8) | 1.79E-207 |
| Missing | 342 (4.0%) | 15,292 (12%) |  |
| Cancer | 1,782 (22) | 15,136 (13) | 3.47E-115 |
| Missing | 342 (4.0%) | 15,292 (12%) |  |
| Charlson Comorbidity Index | 1.00 (0.00-2.00) | 0.00 (0.00-1.00) | <2.2E-16 |
| Missing | 342 (4.0%) | 15,292 (12%) |  |
| Admission method |  |  | 8.12E-224 |
| Elective | 5,242 (62) | 101,732 (77) |  |
| Emergency | 2,899 (34) | 27,690 (21) |  |
| Maternity | 1 (<0.1) | 112 (<0.1) |  |
| Not known | 1 (<0.1) | 27 (<0.1) |  |
| Transfer | 329 (3.9) | 2,530 (1.9) |  |
| Operative category |  |  | 1.86E-155 |
| Inclusive/complex | 50 (0.6) | 384 (0.3) |  |
| Inclusive/major | 4 (<0.1) | 129 (<0.1) |  |
| Intermediate/complex | 348 (4.1) | 6,404 (4.8) |  |
| Intermediate/major | 555 (6.6) | 18,114 (14) |  |
| NOC/complex | 5 (<0.1) | 78 (<0.1) |  |
| Restrictive/complex | 666 (7.9) | 5,618 (4.3) |  |
| Restrictive/major | 1,637 (19) | 31,194 (24) |  |
| Restrictive/NOC | 5,207 (61) | 70,170 (53) |  |
| Surgical specialty |  |  | <2.2E-16 |
| Cardiothoracics | 1,158 (14) | 4,211 (3.2) |  |
| General surgery | 2,229 (26) | 24,604 (19) |  |
| Neurosurgery | 371 (4.4) | 4,902 (3.7) |  |
| Other | 1,865 (22) | 34,937 (26) |  |
| Trauma and orthopaedics | 1,986 (23) | 51,927 (39) |  |
| Urology | 624 (7.4) | 10,080 (7.6) |  |
| Vascular | 239 (2.8) | 1,430 (1.1) |  |
| *^1^* Mean (SD); n (%); Median (Q1, Q3); Median (Q1-Q3) | | | |
| *^2^* p-values reported as <2.2E-16 were below estimable limit of the relevant R function. | | | |

**Supplementary table 2** Participant characteristics – atrial fibrillation

| **Characteristic** | **Controls** | **Cases** | **p-value*^2^*** |
| --- | --- | --- | --- |
|  | N = 135,878*^1^* | N = 2,739*^1^* |  |
| Age | 64 (8) | 72 (6) | <2.2E-16 |
| Sex |  |  | 6.06E-105 |
| Female | 76,143 (56) | 963 (35) |  |
| Male | 59,735 (44) | 1,776 (65) |  |
| BMI (kg/m2) | 27.5 (24.8, 30.9) | 28.4 (25.4, 31.6) | 9.66E-16 |
| Missing | 707 (0.5%) | 12 (0.4%) |  |
| Ethnicity |  |  | 9.49E-07 |
| Asian or Asian British | 2,262 (1.7) | 33 (1.2) |  |
| Black or Black British | 1,646 (1.2) | 8 (0.3) |  |
| Mixed | 669 (0.5) | 6 (0.2) |  |
| Other ethnic group | 1,206 (0.9) | 11 (0.4) |  |
| White | 129,473 (96) | 2,669 (98) |  |
| Missing | 622 (0.5%) | 12 (0.4%) |  |
| Townsend deprivation index | -2.1 (-3.6, 0.6) | -2.1 (-3.6, 0.6) | 0.5739473 |
| Missing | 156 (0.1%) | 0 (0%) |  |
| Alcohol intake |  |  | 1.72E-05 |
| Daily or almost daily | 27,136 (20) | 649 (24) |  |
| Never | 12,180 (9.0) | 210 (7.7) |  |
| Once or twice a week | 34,472 (25) | 677 (25) |  |
| One to three times a month | 15,261 (11) | 268 (9.8) |  |
| Special occasions only | 17,416 (13) | 324 (12) |  |
| Three or four times a week | 29,107 (21) | 606 (22) |  |
| Missing | 306 (0.2%) | 5 (0.2%) |  |
| Smoking status |  |  | 1.86E-09 |
| Current | 14,860 (11) | 266 (9.7) |  |
| Never | 68,370 (50) | 1,243 (45) |  |
| Prefer not to answer | 626 (0.5) | 14 (0.5) |  |
| Previous | 51,882 (38) | 1,215 (44) |  |
| Missing | 140 (0.1%) | 1 (<0.1%) |  |
| Physical activity (days/week) | 4.00 (2.00, 6.00) | 4.00 (2.00, 6.00) | 4.76E-04 |
| Missing | 8,623 (6.3%) | 149 (5.4%) |  |
| Myocardial infarction | 5,975 (5.0) | 406 (15) | 1.88E-122 |
| Missing | 15,710 (12%) | 70 (2.6%) |  |
| Congestive cardiac failure | 2,664 (2.2) | 313 (12) | 2.66E-218 |
| Missing | 15,710 (12%) | 70 (2.6%) |  |
| Cerebrovascular disease | 4,283 (3.6) | 266 (10.0) | 7.73E-67 |
| Missing | 15,710 (12%) | 70 (2.6%) |  |
| Dementia | 426 (0.4) | 23 (0.9) | 3.58E-05 |
| Missing | 15,710 (12%) | 70 (2.6%) |  |
| Chronic pulmonary disease | 14,932 (12) | 543 (20) | 4.80E-34 |
| Missing | 15,710 (12%) | 70 (2.6%) |  |
| Diabetes | 9,134 (7.6) | 359 (13) | 6.63E-29 |
| Missing | 15,710 (12%) | 70 (2.6%) |  |
| Diabetes with complications | 1,216 (1.0) | 64 (2.4) | 6.07E-12 |
| Missing | 15,710 (12%) | 70 (2.6%) |  |
| Renal disease | 3,353 (2.8) | 236 (8.8) | 7.70E-75 |
| Missing | 15,710 (12%) | 70 (2.6%) |  |
| Cancer | 15,829 (13) | 475 (18) | 4.03E-12 |
| Missing | 15,710 (12%) | 70 (2.6%) |  |
| Charlson Comorbidity Index | 0.00 (0.00-1.00) | 1.00 (0.00-2.00) | 8.50E-108 |
| Missing | 15,710 (12%) | 70 (2.6%) |  |
| Admission method |  |  | 2.93E-33 |
| Elective | 104,248 (77) | 1,872 (68) |  |
| Emergency | 28,869 (21) | 745 (27) |  |
| Maternity | 113 (<0.1) | 0 (0) |  |
| Not known | 28 (<0.1) | 0 (0) |  |
| Transfer | 2,620 (1.9) | 122 (4.5) |  |
| Operative category |  |  | 1.26E-59 |
| Inclusive/complex | 389 (0.3) | 34 (1.2) |  |
| Inclusive/major | 132 (<0.1) | 0 (0) |  |
| Intermediate/complex | 6,498 (4.8) | 157 (5.7) |  |
| Intermediate/major | 18,329 (13) | 205 (7.5) |  |
| NOC/complex | 77 (<0.1) | 1 (<0.1) |  |
| Restrictive/complex | 5,948 (4.4) | 151 (5.5) |  |
| Restrictive/major | 32,000 (24) | 435 (16) |  |
| Restrictive/NOC | 72,505 (53) | 1,756 (64) |  |
| Surgical specialty |  |  | <2.2E-16 |
| Cardiothoracics | 4,385 (3.2) | 659 (24) |  |
| General surgery | 26,031 (19) | 324 (12) |  |
| Neurosurgery | 5,150 (3.8) | 54 (2.0) |  |
| Other | 35,563 (26) | 589 (22) |  |
| Trauma and orthopaedics | 52,836 (39) | 894 (33) |  |
| Urology | 10,383 (7.6) | 157 (5.7) |  |
| Vascular | 1,530 (1.1) | 62 (2.3) |  |
| *^1^* Mean (SD); n (%); Median (Q1, Q3); Median (Q1-Q3) | |  |  |
| *^2^* p-values reported as <2.2E-16 were below estimable limit of the relevant R function. | | | |

**Supplementary table 3** Participant characteristics – acute kidney injury

| **Characteristic** | **Controls N = 134,941*^1^*** | **Cases N = 3,002*^1^*** | **p-value*^2^*** |
| --- | --- | --- | --- |
| Age | 64 (8) | 70 (7) | <2.2E-16 |
| Sex |  |  | 6.39E-92 |
| Female | 75,786 (56) | 1,126 (38) |  |
| Male | 59,155 (44) | 1,876 (62) |  |
| BMI (kg/m2) | 27.5 (24.7, 30.8) | 28.9 (25.9, 32.5) | 4.55E-55 |
| Missing | 671 (0.5%) | 22 (0.7%) |  |
| Ethnicity |  |  | 0.26 |
| Asian or Asian British | 2,182 (1.6) | 58 (1.9) |  |
| Black or Black British | 1,561 (1.2) | 46 (1.5) |  |
| Mixed | 654 (0.5) | 13 (0.4) |  |
| Other ethnic group | 1,171 (0.9) | 28 (0.9) |  |
| White | 128,765 (96) | 2,841 (95) |  |
| Missing | 608 (0.5%) | 16 (0.5%) |  |
| Townsend deprivation index | -2.1 (-3.6, 0.6) | -1.9 (-3.6, 1.0) | 0.002 |
| Missing | 152 (0.1%) | 5 (0.2%) |  |
| Alcohol intake |  |  | 2.81E-07 |
| Daily or almost daily | 27,080 (20) | 649 (22) |  |
| Never | 11,895 (8.8) | 326 (11) |  |
| Once or twice a week | 34,263 (25) | 741 (25) |  |
| One to three times a month | 15,112 (11) | 339 (11) |  |
| Prefer not to answer | 156 (0.1) | 7 (0.2) |  |
| Special occasions only | 17,144 (13) | 399 (13) |  |
| Three or four times a week | 29,155 (22) | 537 (18) |  |
| Missing | 136 (0.1%) | 4 (0.1%) |  |
| Smoking status |  |  | 3.25E-26 |
| Current | 14,449 (11) | 466 (16) |  |
| Never | 68,213 (51) | 1,257 (42) |  |
| Prefer not to answer | 607 (0.5) | 22 (0.7) |  |
| Previous | 51,537 (38) | 1,253 (42) |  |
| Missing | 135 (0.1%) | 4 (0.1%) |  |
| Physical activity (days/week) | 4.00 (2.00, 6.00) | 3.00 (2.00, 5.00) | 0.01 |
| Missing | 8,432 (6.2%) | 240 (8.0%) |  |
| Myocardial infarction | 5,863 (4.9) | 365 (13) | 1.16E-76 |
| Missing | 15,651 (12%) | 104 (3.5%) |  |
| Congestive cardiac failure | 2,622 (2.2) | 299 (10) | 4.27E-175 |
| Missing | 15,651 (12%) | 104 (3.5%) |  |
| Cerebrovascular disease | 4,257 (3.6) | 233 (8.0) | 2.35E-36 |
| Missing | 15,651 (12%) | 104 (3.5%) |  |
| Dementia | 391 (0.3) | 42 (1.4) | 5.06E-23 |
| Missing | 15,651 (12%) | 104 (3.5%) |  |
| Chronic pulmonary disease | 14,642 (12) | 631 (22) | 1.65E-52 |
| Missing | 15,651 (12%) | 104 (3.5%) |  |
| Diabetes | 8,704 (7.3) | 538 (19) | 2.12E-113 |
| Missing | 15,651 (12%) | 104 (3.5%) |  |
| Diabetes with complications | 1,012 (0.8) | 111 (3.8) | 2.53E-61 |
| Missing | 15,651 (12%) | 104 (3.5%) |  |
| Renal disease | 2,689 (2.3) | 386 (13) | 4.76E-308 |
| Missing | 15,651 (12%) | 104 (3.5%) |  |
| Cancer | 15,518 (13) | 638 (22) | 3.06E-45 |
| Missing | 15,651 (12%) | 104 (3.5%) |  |
| Charlson Comorbidity Index | 0.00 (0.00-1.00) | 1.00 (0.00-2.00) | 2.64E-284 |
| Missing | 15,651 (12%) | 104 (3.5%) |  |
| Admission method |  |  | 3.70E-198 |
| Elective | 104,168 (77) | 1,610 (54) |  |
| Emergency | 28,071 (21) | 1,281 (43) |  |
| Maternity | 111 (<0.1) | 1 (<0.1) |  |
| Not known | 29 (<0.1) | 0 (0) |  |
| Transfer | 2,562 (1.9) | 110 (3.7) |  |
| Operative category |  |  | 1.14E-36 |
| Inclusive/complex | 379 (0.3) | 11 (0.4) |  |
| Inclusive/major | 130 (<0.1) | 3 (<0.1) |  |
| Intermediate/complex | 6,512 (4.8) | 118 (3.9) |  |
| Intermediate/major | 18,059 (13) | 247 (8.2) |  |
| NOC/complex | 76 (<0.1) | 2 (<0.1) |  |
| Restrictive/complex | 5,874 (4.4) | 218 (7.3) |  |
| Restrictive/major | 31,813 (24) | 560 (19) |  |
| Restrictive/NOC | 72,098 (53) | 1,843 (61) |  |
| Surgical specialty |  |  | 1.49E-220 |
| Cardiothoracics | 4,740 (3.5) | 377 (13) |  |
| General surgery | 25,533 (19) | 734 (24) |  |
| Neurosurgery | 5,114 (3.8) | 76 (2.5) |  |
| Other | 35,281 (26) | 629 (21) |  |
| Trauma and orthopaedics | 52,792 (39) | 760 (25) |  |
| Urology | 10,003 (7.4) | 334 (11) |  |
| Vascular | 1,478 (1.1) | 92 (3.1) |  |
| *^1^* Mean (SD); n (%); Median (Q1, Q3); Median (Q1-Q3) | | | |
| *^2^* p-values reported as <2.2E-16 were below estimable limit of the relevant R function. | | | |

**Supplementary table 4** Participant characteristics – acute myocardial infarction

| **Characteristic** | **Controls N = 123,339*^1^*** | **Cases N = 282*^1^*** | **p-value** |
| --- | --- | --- | --- |
| Age | 64 (8) | 69 (7) | 1.61E-26 |
| Sex |  |  |  |
| Female | 73,106 (59) | 109 (39) | 3.00E-12 |
| Male | 50,233 (41) | 173 (61) |  |
| BMI (kg/m2) | 27.5 (24.7, 30.9) | 27.9 (24.6, 31.5) | 0.31 |
| Missing | 625 (0.5%) | 1 (0.4%) |  |
| Ethnicity |  |  | 0.68 |
| Asian or Asian British | 1,738 (1.4) | 5 (1.8) |  |
| Other ethnic group | 3,787 (3.1) | 8 (2.8) |  |
| White | 117,814 (96) | 269 (95) |  |
| Townsend deprivation index | -2.1 (-3.6, 0.6) | -1.9 (-3.4, 1.1) | 0.14 |
| Missing | 146 (0.1%) | 0 (0%) |  |
| Alcohol intake |  |  | 0.95 |
| Daily or almost daily | 24,522 (20) | 59 (21) |  |
| Never | 10,893 (8.8) | 27 (9.6) |  |
| Once or twice a week | 31,321 (25) | 66 (23) |  |
| One to three times a month | 13,906 (11) | 35 (12) |  |
| Prefer not to answer | 148 (0.1) | 0 (0) |  |
| Special occasions only | 16,000 (13) | 38 (13) |  |
| Three or four times a week | 26,426 (21) | 57 (20) |  |
| Missing | 123 (<0.1%) | 0 (0%) |  |
| Smoking status |  |  | 1.02E-12 |
| Current | 12,880 (11) | 65 (23) |  |
| Never | 63,129 (51) | 98 (35) |  |
| Previous | 46,642 (38) | 117 (42) |  |
| Missing | 688 (0.6%) | 2 (0.7%) |  |
| Physical activity (days/week) | 4.00 (2.00, 6.00) | 3.00 (1.00, 6.00) | 0.24 |
| Missing | 7,851 (6.4%) | 17 (6.0%) |  |
| Myocardial infarction | 1,224 (1.1) | 31 (12) | 1.74E-57 |
| Missing | 14,923 (12%) | 22 (7.8%) |  |
| Congestive cardiac failure | 1,597 (1.5) | 25 (9.6) | 4.62E-26 |
| Missing | 14,923 (12%) | 22 (7.8%) |  |
| Cerebrovascular disease | 3,733 (3.4) | 21 (8.1) | 8.99E-05 |
| Missing | 14,923 (12%) | 22 (7.8%) |  |
| Dementia | 427 (0.4) | 3 (1.2) | 0.15 |
| Missing | 14,923 (12%) | 22 (7.8%) |  |
| Chronic pulmonary disease | 13,345 (12) | 64 (25) | 2.99E-09 |
| Missing | 14,923 (12%) | 22 (7.8%) |  |
| Diabetes | 7,425 (6.8) | 32 (12) | 7.94E-04 |
| Missing | 14,923 (12%) | 22 (7.8%) |  |
| Diabetes with complications | 917 (0.8) | 19 (7.3) | 8.61E-28 |
| Missing | 14,923 (12%) | 22 (7.8%) |  |
| Renal disease | 2,825 (2.6) | 32 (12) | 1.05E-21 |
| Missing | 14,923 (12%) | 22 (7.8%) |  |
| Cancer | 15,166 (14) | 55 (21) | 0.001 |
| Missing | 14,923 (12%) | 22 (7.8%) |  |
| Charlson Comorbidity Index | 0.00 (0.00-1.00) | 1.00 (0.00-2.00) | 5.53E-19 |
| Missing | 14,923 (12%) | 22 (7.8%) |  |
| Admission method |  |  | 2.71E-58 |
| Elective | 98,726 (80) | 115 (41) |  |
| Emergency | 23,434 (19) | 158 (56) |  |
| Maternity | 111 (<0.1) | 0 (0) |  |
| Not known | 27 (<0.1) | 0 (0) |  |
| Transfer | 1,041 (0.8) | 9 (3.2) |  |
| Operative category |  |  | 1.29E-298 |
| Inclusive/complex | 12 (<0.1) | 3 (1.1) |  |
| Inclusive/major | 106 (<0.1) | 0 (0) |  |
| Intermediate/complex | 753 (0.6) | 47 (17) |  |
| Intermediate/major | 16,728 (14) | 16 (5.7) |  |
| NOC/complex | 79 (<0.1) | 0 (0) |  |
| Restrictive/complex | 5,369 (4.4) | 13 (4.6) |  |
| Restrictive/major | 31,643 (26) | 45 (16) |  |
| Restrictive/NOC | 68,649 (56) | 158 (56) |  |
| Surgical specialty |  |  | 5.53E-26 |
| Cardiothoracics | 13 (<0.1) | 0 (0) |  |
| General surgery | 26,219 (21) | 70 (25) |  |
| Neurosurgery | 5,188 (4.2) | 7 (2.5) |  |
| Other | 26,542 (22) | 102 (36) |  |
| Trauma and orthopaedics | 53,384 (43) | 70 (25) |  |
| Urology | 10,488 (8.5) | 12 (4.3) |  |
| Vascular | 1,505 (1.2) | 21 (7.4) |  |
| *^1^* Mean (SD); n (%); Median (Q1, Q3); Median (Q1-Q3) | | | |

**Supplementary table 5** Participant characteristics – stroke

| **Characteristic** | **Controls N = 137,842*^1^*** | **Cases N = 569*^1^*** | **p-value** |
| --- | --- | --- | --- |
| Age | 64 (8) | 67 (7) | 9.77E-23 |
| Sex |  |  | 6.41E-08 |
| Female | 76,725 (56) | 252 (44) |  |
| Male | 61,117 (44) | 317 (56) |  |
| BMI (kg/m2) | 27.5 (24.8, 30.9) | 27.2 (24.6, 30.1) | 0.02 |
| Missing | 692 (0.5%) | 2 (0.4%) |  |
| Ethnicity |  |  | 0.3 |
| Asian or Asian British | 2,267 (1.6) | 10 (1.8) |  |
| Black or Black British | 1,625 (1.2) | 9 (1.6) |  |
| Mixed | 662 (0.5) | 5 (0.9) |  |
| Other ethnic group | 1,850 (1.3) | 6 (1.1) |  |
| White | 131,438 (95) | 539 (95) |  |
| Townsend deprivation index | -2.1 (-3.6, 0.6) | -1.9 (-3.6, 0.4) | 0.48 |
| Missing | 157 (0.1%) | 1 (0.2%) |  |
| Alcohol intake |  |  | 0.16 |
| Daily or almost daily | 27,704 (20) | 136 (24) |  |
| Never | 12,223 (8.9) | 52 (9.2) |  |
| Once or twice a week | 34,922 (25) | 150 (26) |  |
| One to three times a month | 15,433 (11) | 63 (11) |  |
| Special occasions only | 17,601 (13) | 66 (12) |  |
| Three or four times a week | 29,657 (22) | 100 (18) |  |
| Missing | 302 (0.2%) | 2 (0.4%) |  |
| Smoking status |  |  | 8.76E-06 |
| Current | 14,852 (11) | 89 (16) |  |
| Never | 69,408 (50) | 245 (43) |  |
| Prefer not to answer | 630 (0.5) | 7 (1.2) |  |
| Previous | 52,813 (38) | 227 (40) |  |
| Missing | 139 (0.1%) | 1 (0.2%) |  |
| Physical activity (days/week) | 4.00 (2.00, 6.00) | 4.00 (2.00, 6.00) | 0.89 |
| Missing | 8,673 (6.3%) | 39 (6.9%) |  |
| Myocardial infarction | 6,218 (5.1) | 59 (11) | 5.38E-10 |
| Missing | 15,815 (11%) | 40 (7.0%) |  |
| Congestive cardiac failure | 3,099 (2.5) | 42 (7.9) | 1.31E-14 |
| Missing | 15,815 (11%) | 40 (7.0%) |  |
| Cerebrovascular disease | 2,640 (2.2) | 60 (11) | 8.79E-46 |
| Missing | 15,815 (11%) | 40 (7.0%) |  |
| Dementia | 443 (0.4) | 3 (0.6) | 0.68 |
| Missing | 15,815 (11%) | 40 (7.0%) |  |
| Chronic pulmonary disease | 15,348 (13) | 101 (19) | 9.03E-06 |
| Missing | 15,815 (11%) | 40 (7.0%) |  |
| Diabetes | 9,328 (7.6) | 64 (12) | 1.69E-04 |
| Missing | 15,815 (11%) | 40 (7.0%) |  |
| Diabetes with complications | 1,223 (1.0) | 9 (1.7) | 0.16 |
| Missing | 15,815 (11%) | 40 (7.0%) |  |
| Renal disease | 3,521 (2.9) | 43 (8.1) | 2.04E-12 |
| Missing | 15,815 (11%) | 40 (7.0%) |  |
| Cancer | 16,163 (13) | 85 (16) | 0.06 |
| Missing | 15,815 (11%) | 40 (7.0%) |  |
| Charlson Comorbidity Index | 0.00 (0.00-1.00) | 0.00 (0.00-2.00) | 8.83E-16 |
| Missing | 15,815 (11%) | 40 (7.0%) |  |
| Admission method |  |  | 2.28E-114 |
| Elective | 105,902 (77) | 215 (38) |  |
| Emergency | 29,126 (21) | 302 (53) |  |
| Maternity | 113 (<0.1) | 0 (0) |  |
| Not known | 29 (<0.1) | 0 (0) |  |
| Transfer | 2,672 (1.9) | 52 (9.1) |  |
| Operative category |  |  | 1.26E-33 |
| Inclusive/complex | 400 (0.3) | 6 (1.1) |  |
| Inclusive/major | 133 (<0.1) | 1 (0.2) |  |
| Intermediate/complex | 6,635 (4.8) | 32 (5.6) |  |
| Intermediate/major | 18,566 (13) | 22 (3.9) |  |
| NOC/complex | 78 (<0.1) | 1 (0.2) |  |
| Restrictive/complex | 6,011 (4.4) | 57 (10) |  |
| Restrictive/major | 32,444 (24) | 53 (9.3) |  |
| Restrictive/NOC | 73,575 (53) | 397 (70) |  |
| Surgical specialty |  |  | 4.20E-285 |
| Cardiothoracics | 4,985 (3.6) | 103 (18) |  |
| General surgery | 26,343 (19) | 54 (9.5) |  |
| Neurosurgery | 4,979 (3.6) | 151 (27) |  |
| Other | 36,064 (26) | 154 (27) |  |
| Trauma and orthopaedics | 53,501 (39) | 65 (11) |  |
| Urology | 10,543 (7.6) | 18 (3.2) |  |
| Vascular | 1,427 (1.0) | 24 (4.2) |  |
| *^1^* Mean (SD); n (%); Median (Q1, Q3); Median (Q1-Q3) | | | |

**Supplementary table 6** Participant characteristics – surgical site infection

| **Characteristic** | **Controls N = 136,120*^1^*** | **Cases N = 2,701*^1^*** | **p-value*^2^*** |
| --- | --- | --- | --- |
| Age | 64 (8) | 65 (8) | 0.01 |
| Sex |  |  | 9.81E-07 |
| Female | 75,555 (56) | 1,371 (51) |  |
| Male | 60,565 (44) | 1,330 (49) |  |
| BMI (kg/m2) | 27.5 (24.8, 30.8) | 28.6 (25.5, 32.3) | 1.37E-26 |
| Missing | 680 (0.5%) | 17 (0.6%) |  |
| Ethnicity |  |  | 0.23 |
| Asian or Asian British | 2,226 (1.6) | 54 (2.0) |  |
| Black or Black British | 1,596 (1.2) | 38 (1.4) |  |
| Mixed | 656 (0.5) | 12 (0.4) |  |
| Other ethnic group | 1,183 (0.9) | 29 (1.1) |  |
| White | 129,839 (96) | 2,557 (95) |  |
| Missing | 620 (0.5%) | 11 (0.4%) |  |
| Townsend deprivation index | -2.1 (-3.6, 0.6) | -1.8 (-3.5, 1.3) | 5.64E-08 |
| Missing | 156 (0.1%) | 2 (<0.1%) |  |
| Alcohol intake |  |  | 8.27E-04 |
| Daily or almost daily | 27,413 (20) | 558 (21) |  |
| Never | 12,044 (8.9) | 274 (10) |  |
| Once or twice a week | 34,461 (25) | 714 (26) |  |
| One to three times a month | 15,206 (11) | 289 (11) |  |
| Prefer not to answer | 162 (0.1) | 6 (0.2) |  |
| Special occasions only | 17,338 (13) | 360 (13) |  |
| Three or four times a week | 29,358 (22) | 496 (18) |  |
| Missing | 138 (0.1%) | 4 (0.1%) |  |
| Smoking status |  |  | 9.06E-16 |
| Current | 14,637 (11) | 404 (15) |  |
| Never | 68,629 (50) | 1,180 (44) |  |
| Prefer not to answer | 624 (0.5) | 18 (0.7) |  |
| Previous | 52,093 (38) | 1,095 (41) |  |
| Missing | 137 (0.1%) | 4 (0.1%) |  |
| Physical activity (days/week) | 4.00 (2.00, 6.00) | 3.00 (2.00, 6.00) | 0.06244312 |
| Missing | 8,574 (6.3%) | 176 (6.5%) |  |
| Myocardial infarction | 6,253 (5.2) | 197 (7.6) | 4.93E-08 |
| Missing | 15,688 (12%) | 119 (4.4%) |  |
| Congestive cardiac failure | 3,136 (2.6) | 128 (5.0) | 2.87E-13 |
| Missing | 15,688 (12%) | 119 (4.4%) |  |
| Cerebrovascular disease | 4,574 (3.8) | 137 (5.3) | 9.67E-05 |
| Missing | 15,688 (12%) | 119 (4.4%) |  |
| Dementia | 475 (0.4) | 12 (0.5) | 0.68560093 |
| Missing | 15,688 (12%) | 119 (4.4%) |  |
| Chronic pulmonary disease | 15,006 (12) | 521 (20) | 2.18E-31 |
| Missing | 15,688 (12%) | 119 (4.4%) |  |
| Diabetes | 9,212 (7.6) | 353 (14) | 1.84E-29 |
| Missing | 15,688 (12%) | 119 (4.4%) |  |
| Diabetes with complications | 1,222 (1.0) | 70 (2.7) | 1.35E-16 |
| Missing | 15,688 (12%) | 119 (4.4%) |  |
| Renal disease | 3,553 (3.0) | 147 (5.7) | 1.09E-15 |
| Missing | 15,688 (12%) | 119 (4.4%) |  |
| Cancer | 15,587 (13) | 715 (28) | 9.36E-106 |
| Missing | 15,688 (12%) | 119 (4.4%) |  |
| Charlson Comorbidity Index | 0.00 (0.00-1.00) | 1.00 (0.00-2.00) | 9.59E-201 |
| Missing | 15,688 (12%) | 119 (4.4%) |  |
| Admission method |  |  | 6.48E-20 |
| Elective | 104,191 (77) | 1,855 (69) |  |
| Emergency | 29,088 (21) | 765 (28) |  |
| Maternity | 111 (<0.1) | 0 (0) |  |
| Not known | 28 (<0.1) | 1 (<0.1) |  |
| Transfer | 2,702 (2.0) | 80 (3.0) |  |
| Operative category |  |  | 4.83E-119 |
| Inclusive/complex | 409 (0.3) | 2 (<0.1) |  |
| Inclusive/major | 131 (<0.1) | 1 (<0.1) |  |
| Intermediate/complex | 6,682 (4.9) | 16 (0.6) |  |
| Intermediate/major | 18,440 (14) | 90 (3.3) |  |
| NOC/complex | 80 (<0.1) | 1 (<0.1) |  |
| Restrictive/complex | 5,859 (4.3) | 283 (10) |  |
| Restrictive/major | 31,867 (23) | 676 (25) |  |
| Restrictive/NOC | 72,652 (53) | 1,632 (60) |  |
| Surgical specialty |  |  | <2.2E-16 |
| Cardiothoracics | 4,946 (3.6) | 195 (7.2) |  |
| General surgery | 25,165 (18) | 1,288 (48) |  |
| Neurosurgery | 5,082 (3.7) | 116 (4.3) |  |
| Other | 35,859 (26) | 548 (20) |  |
| Trauma and orthopaedics | 53,113 (39) | 334 (12) |  |
| Urology | 10,405 (7.6) | 147 (5.4) |  |
| Vascular | 1,550 (1.1) | 73 (2.7) |  |
| *^1^* Mean (SD); n (%); Median (Q1, Q3); Median (Q1-Q3) | | | |
| *^2^* p-values reported as <2.2E-16 were below estimable limit of the relevant R function. | | | |

**Supplementary table 7** Mapped genes from GWAS of postoperative atrial fibrillation

| chr | symbol | type | minGwasP | IndSigSNPs |
| --- | --- | --- | --- | --- |
| 4 | PITX2 | protein_coding | 1.05E-19 | 4:111697815_TG_T;rs200657990:4:111697815_TG_T |
| 4 | RP11-777N19.1 | lincRNA | 1.05E-19 | 4:111697815_TG_T;rs200657990;rs723363 |
| 6 | ERMARD | protein_coding | 3.08E-13 | 4:111697815_TG_T;rs200657990 |
| 4 | AC004067.5 | antisense | 3.54E-10 | 4:111607460_ATT_A |
| 16 | SPSB3 | protein_coding | 2.32E-08 | rs145672192 |
| 16 | MSRB1 | protein_coding | 2.32E-08 | rs145672192 |
| 16 | AC005363.9 | pseudogene | 2.32E-08 | rs145672192 |
| 16 | NPW | protein_coding | 2.32E-08 | rs145672192 |
| 16 | NTHL1 | protein_coding | 2.32E-08 | rs145672192 |
| 16 | TSC2 | protein_coding | 2.32E-08 | rs145672192 |
| 16 | TRAF7 | protein_coding | 2.32E-08 | rs145672192 |
| 16 | CASKIN1 | protein_coding | 2.32E-08 | rs145672192 |
| 16 | ECI1 | protein_coding | 2.32E-08 | rs145672192 |
| 16 | CLDN9 | protein_coding | 2.32E-08 | rs145672192 |
| 16 | CLDN6 | protein_coding | 2.32E-08 | rs145672192 |
| 16 | TNFRSF12A | protein_coding | 2.32E-08 | rs145672192 |
| 16 | HCFC1R1 | protein_coding | 2.32E-08 | rs145672192 |
| 16 | THOC6 | protein_coding | 2.32E-08 | rs145672192 |
| 1 | KCNN3 | protein_coding | 4.08E-08 | rs36088503 |
| 1 | PBXIP1 | protein_coding | 4.08E-08 | rs36088503 |
| 1 | PYGO2 | protein_coding | 4.08E-08 | rs36088503 |
| 1 | SHC1 | protein_coding | 4.08E-08 | rs36088503 |
| 1 | RP11-307C12.11 | antisense | 4.08E-08 | rs36088503 |
| 1 | ADAM15 | protein_coding | 4.08E-08 | rs36088503 |
| 1 | MUC1 | protein_coding | 4.08E-08 | rs36088503 |
| 1 | THBS3 | protein_coding | 4.08E-08 | rs36088503 |
| 4 | EGF | protein_coding | 4.35E-08 | rs723363;rs4571403 |
| 1 | GBAP1 | pseudogene | 6.68E-08 | rs36088503 |
| 1 | SHE | protein_coding | 4.18E-07 | rs36088503 |
| 1 | TDRD10 | protein_coding | 4.18E-07 | rs36088503 |
| 1 | UBE2Q1 | protein_coding | 4.18E-07 | rs36088503 |
| 1 | CHRNB2 | protein_coding | 4.18E-07 | rs36088503 |
| 1 | ADAR | protein_coding | 4.18E-07 | rs36088503 |
| 1 | PMVK | protein_coding | 4.18E-07 | rs36088503 |
| 1 | RP11-307C12.12 | antisense | 4.18E-07 | rs36088503 |
| 1 | CKS1B | protein_coding | 4.18E-07 | rs36088503 |
| 1 | FLAD1 | protein_coding | 4.18E-07 | rs36088503 |
| 1 | ZBTB7B | protein_coding | 4.18E-07 | rs36088503 |
| 1 | DPM3 | protein_coding | 4.18E-07 | rs36088503 |
| 1 | GBA | protein_coding | 4.18E-07 | rs36088503 |
| 1 | MSTO2P | pseudogene | 4.83E-06 | rs36088503 |
| 16 | CCDC154 | protein_coding | 6.40E-06 | rs145672192 |

**Supplementary table 8** Independent significant SNPs from GWAS of postoperative atrial fibrillation with 180 day control washout period

| **rsID*** | **Chr:Pos**** | **Nearest gene** | **Function** | **Effect allele** | **OR (95% CI)** | **p** |
| --- | --- | --- | --- | --- | --- | --- |
| - | 4:111697815_TG_T | RP11-777N19.1 | intergenic | TG | 0.73 (0.68-0.78) | 6.08E-20 |
| rs200657990 | 4:111679136 | RP11-777N19.1 | intergenic | A | 0.71 (0.65-0.77) | 1.24E-13 |
| rs10032150 | 4:111646618 | RP11-777N19.1 | intergenic | A | 0.81 (0.76-0.86) | 8.83E-11 |
| - | 4:111607460_ATT_A | PITX2 | intergenic | ATT | 0.78 (0.73-0.84) | 2.74E-10 |
| rs72667982 | 4:111673642 | RP11-777N19.1 | intergenic | A | 0.81 (0.76-0.87) | 2.48E-09 |
| rs145672192 | 16:2227985 | TRAF7:CASKIN1 | UTR3 | G | 2.33 (1.67-3.25) | 2.04E-08 |
| rs4571403 | 4:111613368 | PITX2 | intergenic | G | 1.19 (1.12-1.26) | 3.09E-08 |
| rs723363 | 4:111724501 | RP11-777N19.1 | intergenic | T | 1.18 (1.11-1.25) | 4.21E-08 |

**Supplementary table 9** Independent significant SNPs from GWAS of postoperative atrial fibrillation adjusted for surgical specialty

| **rsID*** | **Chr:Pos**** | **Nearest gene** | **Function** | **Effect allele** | **OR (95% CI)** | **p** |
| --- | --- | --- | --- | --- | --- | --- |
| - | 4:111697815_TG_T | *RP11-777N19.1* | intergenic | TG | 0.72 (0.68-0.77) | 4.14E-20 |
| rs200657990 | 4:111679136 | *RP11-777N19.1* | intergenic | A | 0.7 (0.64-0.76) | 4.47E-14 |
| rs10032150 | 4:111646618 | *RP11-777N19.1* | intergenic | A | 0.81 (0.76-0.86) | 1.29E-10 |
| rs72667982 | 4:111673642 | *RP11-777N19.1* | intergenic | A | 0.8 (0.74-0.85) | 1.74E-10 |
| - | 4:111607460_ATT_A | *PITX2* | intergenic | ATT | 0.79 (0.73-0.85) | 1.04E-09 |
| rs723363 | 4:111724501 | *RP11-777N19.1* | intergenic | T | 1.19 (1.12-1.26) | 1.96E-08 |
| rs1218566 | 1 | KCNN3 | intronic | T | 1.18 (1.11-1.25) | 3.08E-08 |
| rs36088503 | 1:154812943 | *KCNN3* | intronic | G | 0.82 (0.77-0.88) | 3.87E-08 |

**Supplementary table 10** Independent significant SNPs from GWAS of postoperative atrial fibrillation after non-cardiothoracic surgery

| **rsID** | **Chr** | **Nearest gene** | **Function** | **Effect allele** | **OR (95% CI)** | **p** |
| --- | --- | --- | --- | --- | --- | --- |
| rs6843082 | 4 | *RP11-777N19.1* | ncRNA_exonic | G | 1.41 (1.3-1.52) | 3.81 x 10^-16^ |
| rs1906593 | 4 | *RP11-777N19.1* | intergenic | C | 0.71 (0.64-0.78) | 7.88 x 10^-11^ |
| rs34102391 | 4 | *RP11-777N19.1* | intergenic | C | 1.31 (1.2-1.42) | 3.33 x 10^-10^ |
| rs2595093 | 4 | *PITX2* | intergenic | A | 1.33 (1.22-1.45) | 8.88 x 10^-10^ |
| rs72667982 | 4 | *RP11-777N19.1* | intergenic | A | 0.79 (0.72-0.85) | 1.17 x 10^-8^ |
| rs12052983 | 2 | *PLEKHA3* | intronic | G | 0.8 (0.74-0.86) | 1.51 x 10^-8^ |

**Supplementary table 11** Genotype-by-specialty (G x E) interaction results for postoperative atrial fibrillation primary GWAS lead SNPs

| **SNP** | **Interaction_Beta** | **Interaction_P** | **NonCardiac_Beta** | **NonCardiac_P** |
| --- | --- | --- | --- | --- |
| 4:111697815_TG_T_TG | 0.16 | 0.06 | -0.36 | 8.56E-22 |
| rs145672192_G | -1.07 | 0.007 | 1.13 | 2.08E-06 |
| rs36088503_G | 0.07 | 0.43 | -0.21 | 1.01E-07 |

**Supplementary table 12** Independent SNPs meeting an exploratory p-value threshold of 1 x 10^-6^ for postoperative acute kidney injury (AKI), myocardial infarction (AMI) and surgical site infection (SSI)

| **Outcome** | **rsID** | **Chr** | **Nearest gene** | **Function** | **Effect allele** | **Beta (SE)** | **p** |
| --- | --- | --- | --- | --- | --- | --- | --- |
| AKI | rs117097059 | 10 | *RP11-428L9.2* | Intergenic | C | -0.31 (0.06) | 4.8 x 10^-7^ |
| AKI | rs11256067 | 10 | *RP11-428L9.2* | Intergenic | T | -0.42 (0.08) | 9.7 x 10^-7^ |
| AKI | rs430102 | 12 | *CLLU1OS* | Intergenic | C | 0.14 (0.03) | 5.3 x 10^-7^ |
| AMI | rs150601474 | 7 | *BBS9* | Intronic | G | -1.21 (0.2) | 6.2 x 10^-7^ |
| AMI | rs3743157 | 15 | *PDE8A* | Intronic | C | -0.53 (0.09) | 9.3 x 10^-8^ |
| AMI | rs142068482 | 20 | *CDH4* | Intergenic | T | 2.3 (0.56) | 4.5 x 10^-7^ |
| SSI | rs138492269 | 1 | *RGS5* | Intergenic | A | -0.91 (0.16) | 3.3 x 10^-7^ |
| SSI | rs562534599 | 4 | *CCNG2:RP11-625I7.1* | ncRNA_intronic | C | 2.73 (0.87) | 6.5 x 10^-7^ |
| SSI | rs13190087 | 5 | *TERT* | Intergenic | A | -0.35 (0.07) | 3.7 x 10^-7^ |
| SSI | rs186310511 | 6 | *F13A1* | Intergenic | A | -0.95 (0.16) | 2.6 x 10^-7^ |
| SSI | rs561001924 | 6 | *LINC00336* | Intergenic | C | -0.15 (0.03) | 8.3 x 10^-7^ |
| SSI | rs4930790 | 12 | *ANO2* | Intronic | G | -0.24 (0.05) | 6 x 10^-7^ |
| SSI | rs111784580 | 19 | *SIGLEC21P* | Intergenic | C | -0.44 (0.08) | 1.9 x 10^-7^ |
| SSI | rs62201522 | 20 | *BPI:CTD-2308N23.2* | ncRNA_intronic | T | -0.74 (0.14) | 8.3x 10^-7^ |

**Supplementary table 13** Cross-trait genetic correlation results for all postoperative phenotypes

| **p1** | **p2** | **rg** | **se** | **z** | **p** | **h2_obs** | **h2_obs_se** |
| --- | --- | --- | --- | --- | --- | --- | --- |
| AF | AKI | -0.49 | 0.39 | -1.24 | 0.21 | 0.0092 | 0.003 |
| AF | AMI | NA | NA | NA | NA | NA | NA |
| AF | SSI | 0.75 | 1.58 | 0.48 | 0.63 | 0.0012 | 0.0032 |
| AF | Stroke | -0.64 | 1.21 | -0.53 | 0.6 | 0.0015 | 0.0033 |
| AKI | AMI | 0.32 | 1.08 | 0.29 | 0.77 | 0.0009 | 0.0033 |
| AKI | SSI | 1.09 | 1.18 | 0.92 | 0.36 | 0.0017 | 0.0032 |
| AKI | Stroke | 0.56 | 1.05 | 0.53 | 0.6 | 0.0016 | 0.0033 |
| AMI | SSI | -1.37 | 4.67 | -0.29 | 0.77 | 0.0017 | 0.0032 |
| AMI | Stroke | -0.23 | 3.03 | -0.07 | 0.94 | 0.0016 | 0.0033 |
| SSI | Stroke | -0.13 | 1.64 | -0.08 | 0.94 | 0.0016 | 0.0033 |

p1 = trait 1, p2 = trait 2, rg = genetic correlation, se = standard error of rg, p = p-value for rg; h2_obs, h2_obs_se = observed scale h2 for trait 2 and standard error. NA results are due to the low observed heritability and small sample size.

**Supplementary table 14** Cross-trait genetic correlation results between postoperative (UK Biobank) and ambulatory (FinnGen) phenotypes

| **Postoperative phenotype** | **FinnGen phenotype** | **rg** | **se** | **z** | **p** | **h2_obs** | **h2_obs_se** |
| --- | --- | --- | --- | --- | --- | --- | --- |
| AF | AF | 0.69 | 0.21 | 3.3 | 0.001 | 0.09 | 0.01 |
| AKI | AKI | 0.07 | 0.3 | 0.25 | 0.8 | 0.004 | 0.001 |
| AKI | CKD | 0.42 | 0.22 | 1.93 | 0.054 | 0.006 | 0.001 |
| AMI | AMI | NA | NA | NA | NA | NA | NA |
| Stroke | Stroke | -0.26 | 0.51 | -0.51 | 0.61 | 0.01 | 0.002 |

p1 = trait 1, p2 = trait 2, rg = genetic correlation, se = standard error of rg, p = p-value for rg; h2_obs, h2_obs_se = observed scale h2 for trait 2 and standard error. NA results for AMI, as above

**Supplementary table 15** Polygenic risk scores passing matching

| **Postoperative phenotype** | **PGS ID** | **Reported Trait** | **Number of Variants** | **Publication (doi)** |
| --- | --- | --- | --- | --- |
| AF | PGS000016 | Atrial fibrillation | 6730541 | 10.1038/s41588-018-0183-z |
| AF | PGS000331 | Atrial fibrillation | 6183494 | 10.1038/s41591-020-0800-0 |
| AF | PGS000338 | Atrial fibrillation | 97 | 10.1002/ejhf.1735 |
| AF | PGS000727 | Atrial fibrillation | 2210336 | 10.1038/s42255-021-00478-5 |
| AF | PGS001339 | Atrial fibrillation and flutter (time-to-event) | 2142 | 10.1371/journal.pgen.1010105 |
| AF | PGS001340 | Atrial fibrillation | 2955 | 10.1371/journal.pgen.1010105 |
| AF | PGS001356 | Atrial fibrillation | 2996793 | 10.1161/circgen.120.003128 |
| AF | PGS001841 | Atrial fibrillation and flutter | 3980 | 10.1016/j.ajhg.2021.11.008 |
| AF | PGS002050 | Atrial fibrillation and flutter | 554908 | 10.1016/j.ajhg.2021.11.008 |
| AF | PGS002756 | Atrial fibrillation | 1091491 | 10.1016/j.ajhg.2022.10.009 |
| AF | PGS002773 | Incident atrial fibrillation | 265 | 10.1371/journal.pone.0278764 |
| AF | PGS002774 | Incident atrial fibrillation | 216837 | 10.1371/journal.pone.0278764 |
| AF | PGS002814 | Atrial fibrillation | 4520 | 10.1038/s41588-022-01284-9 |
| AF | PGS003461 | Atrial fibrillation | 162 | 10.1038/s41746-023-00781-3 |
| AF | PGS003761 | Atrial fibrillation | 165 | 10.1186/s12916-023-02798-7 |
| AF | PGS004186 | Atrial fibrillation | 3082 | 10.1038/s41598-023-37580-5 |
| AF | PGS004187 | Atrial fibrillation | 5118 | 10.1038/s41598-023-37580-5 |
| AF | PGS004188 | Atrial fibrillation | 2420 | 10.1038/s41598-023-37580-5 |
| AF | PGS004189 | Atrial fibrillation | 3434 | 10.1038/s41598-023-37580-5 |
| AF | PGS004190 | Atrial fibrillation | 1972 | 10.1038/s41598-023-37580-5 |
| AKI | PGS000708 | Kidney failure | 183272 | 10.1038/s41588-020-00757-z |
| AKI | PGS000728 | Chronic kidney disease | 1958860 | 10.1038/s42255-021-00478-5 |
| AKI | PGS000859 | Chronic kidney disease | 34 | 10.1038/s41588-021-00948-2 |
| AKI | PGS001272 | Chronic renal failure (time-to-event) | 158 | 10.1371/journal.pgen.1010105 |
| AKI | PGS002237 | Chronic kidney disease (stage 3 or greater) | 471316 | 10.1038/s41591-022-01869-1 |
| AKI | PGS002757 | Chronic kidney disease | 1090783 | 10.1016/j.ajhg.2022.10.009 |
| AKI | PGS003988 | Chronic kidney disease (CKD) | 1141637 | 10.1101/2023.11.20.23298215 |
| AKI | PGS004004 | Chronic kidney disease (CKD) | 15373 | 10.1101/2023.11.20.23298215 |
| AKI | PGS004016 | Chronic kidney disease (CKD) | 88605 | 10.1101/2023.11.20.23298215 |
| AKI | PGS004030 | Chronic kidney disease (CKD) | 1050295 | 10.1101/2023.11.20.23298215 |
| AKI | PGS004045 | Chronic kidney disease (CKD) | 1050295 | 10.1101/2023.11.20.23298215 |
| AKI | PGS004058 | Chronic kidney disease (CKD) | 846995 | 10.1101/2023.11.20.23298215 |
| AKI | PGS004074 | Chronic kidney disease (CKD) | 846995 | 10.1101/2023.11.20.23298215 |
| AKI | PGS004088 | Chronic kidney disease (CKD) | 1109217 | 10.1101/2023.11.20.23298215 |
| AKI | PGS004101 | Chronic kidney disease (CKD) | 1109217 | 10.1101/2023.11.20.23298215 |
| AKI | PGS004112 | Chronic kidney disease (CKD) | 301 | 10.1101/2023.11.20.23298215 |
| AKI | PGS004128 | Chronic kidney disease (CKD) | 8543 | 10.1101/2023.11.20.23298215 |
| AKI | PGS004142 | Chronic kidney disease (CKD) | 804867 | 10.1101/2023.11.20.23298215 |
| AKI | PGS004158 | Chronic kidney disease (CKD) | 1135455 | 10.1101/2023.11.20.23298215 |
| AKI | PGS004224 | Chronic kidney disease | 241 | 10.18332/tid/162607 |
| AMI | PGS000710 | Myocardial infarction | 183566 | 10.1038/s41588-020-00757-z |
| AMI | PGS001048 | NSTEMI (algorithmically-defined) | 687 | 10.1371/journal.pgen.1010105 |
| AMI | PGS001314 | Acute myocardial infarction (time-to-event) | 1108 | 10.1371/journal.pgen.1010105 |
| AMI | PGS001315 | Myocardial infarction | 1788 | 10.1371/journal.pgen.1010105 |
| AMI | PGS001316 | Myocardial infarction (algorithmically-defined) | 1831 | 10.1371/journal.pgen.1010105 |
| AMI | PGS001317 | Vascular/heart problems diagnosed by doctor Heart attack | 1030 | 10.1371/journal.pgen.1010105 |
| Stroke | PGS000038 | Stroke | 90 | 10.1136/bmj.k4168 |
| Stroke | PGS000039 | Ischemic stroke | 3225583 | 10.1038/s41467-019-13848-1 |
| Stroke | PGS000665 | Ischemic stroke | 32 | 10.1161/circulationaha.120.051927 |
| Stroke | PGS000911 | Ischemic stroke | 530933 | 10.1161/circgen.120.003168 |
| Stroke | PGS001793 | Stroke | 910099 | 10.1016/j.xgen.2022.100241 |
| Stroke | PGS001798 | Stroke | 884168 | 10.1016/j.xgen.2022.100241 |
| Stroke | PGS002259 | Stroke | 534 | 10.1212/wnl.0000000000012263 |
| Stroke | PGS002724 | Ischemic stroke | 1213574 | 10.1038/s41586-022-05165-3 |
| Stroke | PGS002725 | Ischemic stroke | 6010730 | 10.1038/s41586-022-05165-3 |
| Stroke | PGS002770 | Stroke | 1088719 | 10.1016/j.ajhg.2022.10.009 |
| Stroke | PGS003984 | Stroke | 1121845 | 10.1101/2023.11.20.23298215 |
| Stroke | PGS004000 | Stroke | 2371 | 10.1101/2023.11.20.23298215 |
| Stroke | PGS004015 | Stroke | 65138 | 10.1101/2023.11.20.23298215 |
| Stroke | PGS004026 | Stroke | 1011468 | 10.1101/2023.11.20.23298215 |
| Stroke | PGS004041 | Stroke | 1011468 | 10.1101/2023.11.20.23298215 |
| Stroke | PGS004054 | Stroke | 852173 | 10.1101/2023.11.20.23298215 |
| Stroke | PGS004070 | Stroke | 852173 | 10.1101/2023.11.20.23298215 |
| Stroke | PGS004084 | Stroke | 1091747 | 10.1101/2023.11.20.23298215 |
| Stroke | PGS004098 | Stroke | 1091747 | 10.1101/2023.11.20.23298215 |
| Stroke | PGS004108 | Stroke | 13 | 10.1101/2023.11.20.23298215 |
| Stroke | PGS004124 | Stroke | 5808 | 10.1101/2023.11.20.23298215 |
| Stroke | PGS004138 | Stroke | 888649 | 10.1101/2023.11.20.23298215 |
| Stroke | PGS004154 | Stroke | 1116976 | 10.1101/2023.11.20.23298215 |
| Stroke | PGS004322 | Ischemic stroke | 30 | 10.3389/fcvm.2023.1076745 |
| Stroke | PGS004597 | Ischemic stroke | 32 | 10.3390/nu15040864 |

**Supplementary table 16** Odds ratios for postoperative complication by polygenic risk score (PRS) quintile. Ensemble PRS calculated from all available scores. Results are adjusted for age and sex.

|  | **AF** |  | **AKI** |  | **AMI** |  | **Stroke** |  |
| --- | --- | --- | --- | --- | --- | --- | --- | --- |
| **Term** | **OR**  **(95% CI)** | **P** | **OR**  **(95% CI)** | **P** | **OR**  **(95% CI)** | **P** | **OR**  **(95% CI)** | **P** |
| PRS quintile 1 | 1.00 (reference) | - | 1.00 (reference) | - | 1.00 (reference) | - | 1.00 (reference) | - |
| PRS quintile 2 | 1.50  (1.28–1.77) | <0.001 | 0.99  (0.88–1.12) | 0.87 | 1.38  (0.87–2.19) | 0.17 | 1.26  (0.95–1.69) | 0.11 |
| PRS quintile 3 | 2.11  (1.81–2.46) | <0.001 | 1.00  (0.89–1.13) | 0.94 | 1.68  (1.08–2.64) | 0.02 | 1.23  (0.92–1.65) | 0.02 |
| PRS quintile 4 | 2.76  (2.39–3.21) | <0.001 | 1.17  (1.04–1.32) | 0.007 | 2.24  (1.48–3.46) | <0.001 | 1.53  (1.16–2.02) | 0.003 |
| PRS quintile 5 | 4.92  (4.28–5.67) | <0.001 | 1.23  (1.09–1.37) | <0.001 | 3.81  (2.60–5.74) | <0.001 | 1.84  (1.41–2.41) | <0.001 |
| Age | 1.20  (1.20–1.21) | <0.001 | 1.12  (1.11–1.13) | <0.001 | 1.09  (1.08–1.11) | <0.001 | 1.05  (1.04–1.07) | <0.001 |
| Sex - male | 2.18  (2.02–2.37) | <0.001 | 1.99  (1.84–2.14) | <0.001 | 2.45  (1.94–3.11) | <0.001 | 1.48  (1.26–1.75) | <0.001 |
| AUC  (full model) | 0.83  (0.82–0.84) |  | 0.73  (0.72–0.74) |  | 0.74  (0.71–0.77) |  | 0.63  (0.61–0.66) |  |

**Supplementary table 17** Odds ratios for postoperative complication by polygenic risk score (PRS) quintile. Single score for each outcome based on best reported development performance (AUROC or C-statistic). Results are adjusted for age and sex.

|  | **AF** |  | **AKI** |  | **AMI** |  | **Stroke** |  |
| --- | --- | --- | --- | --- | --- | --- | --- | --- |
| **Term** | **OR**  **(95% CI)** | **P** | **OR**  **(95% CI)** | **P** | **OR**  **(95% CI)** | **P** | **OR**  **(95% CI)** | **P** |
| PRS quintile 1 | 1.00 (reference) | - | 1.00 (reference) | - | 1.00 (reference) | - | 1.00 (reference) | - |
| PRS quintile 2 | 1.20  (1.04–1.39) | 0.01 | 1.02 (0.91–1.15) | 0.73 | 1.03 (0.69–1.53) | 0.89 | 1.09 (0.83–1.45) | 0.52 |
| PRS quintile 3 | 1.55  (1.35–1.78) | <0.001 | 1.09 (0.97–1.23) | 0.15 | 1.07 (0.72–1.59) | 0.74 | 1.19 (0.91–1.57) | 0.2 |
| PRS quintile 4 | 1.83  (1.60–2.09) | <0.001 | 1.10 (0.98–1.23) | 0.12 | 1.34 (0.92–1.96) | 0.13 | 1.22 (0.93–1.60) | 0.15 |
| PRS quintile 5 | 2.66  (2.34–3.02) | <0.001 | 1.16 (1.04–1.31) | 0.01 | 1.86 (1.31–2.68) | <0.001 | 1.37 (1.06–1.79) | 0.02 |
| Age | 1.20  (1.19–1.21) | <0.001 | 1.12 (1.11–1.13) | <0.001 | 1.09 (1.07–1.11) | <0.001 | 1.06 (1.04–1.07) | <0.001 |
| Sex - male | 2.13  (1.97–2.31) | <0.001 | 2.01 (1.87–2.17) | <0.001 | 2.39 (1.88–3.04) | <0.001 | 1.53 (1.30–1.81) | <0.001 |
| AUC  (full model) | 0.82  (0.81–0.82) |  | 0.72  (0.72–0.74) |  | 0.72  (0.69–0.75) |  | 0.63  (0.61–0.65) |  |

**Supplementary table 18** Odds ratios for postoperative complication by polygenic risk score (PRS) quintile. Single score for each outcome based on largest discovery sample size. Results are adjusted for age and sex.

|  | **AF** |  | **AKI** |  | **AMI** |  | **Stroke** |  |
| --- | --- | --- | --- | --- | --- | --- | --- | --- |
| **Term** | **OR**  **(95% CI)** | **P** | **OR**  **(95% CI)** | **P** | **OR**  **(95% CI)** | **P** | **OR**  **(95% CI)** | **P** |
| PRS quintile 1 | 1.00 (reference) | - | 1.00 (reference) | - | 1.00 (reference) | - | 1.00 (reference) | - |
| PRS quintile 2 | 1.44 (1.23–1.69) | <0.001 | 0.99 (0.88–1.12) | 0.93 | 1.53 (1.00–2.38) | 0.05 | 1.09 (0.83–1.45) | 0.52 |
| PRS quintile 3 | 1.72 (1.48–2.01) | <0.001 | 1.08 (0.96–1.22) | 0.19 | 1.46 (0.94–2.29) | 0.09 | 1.19 (0.91–1.57) | 0.2 |
| PRS quintile 4 | 2.33 (2.02–2.70) | <0.001 | 1.14 (1.01–1.28) | 0.03 | 1.59 (1.03–2.47) | 0.04 | 1.22 (0.93–1.60) | 0.15 |
| PRS quintile 5 | 4.01 (3.50–4.60) | <0.001 | 1.28 (1.14–1.44) | <0.001 | 3.45 (2.37–5.14) | <0.001 | 1.37 (1.06–1.79) | 0.02 |
| Age | 1.21 (1.20–1.22) | <0.001 | 1.12 (1.11–1.13) | <0.001 | 1.09 (1.07–1.11) | <0.001 | 1.06 (1.04–1.07) | <0.001 |
| Sex - male | 2.15 (1.98–2.33) | <0.001 | 2.01 (1.87–2.17) | <0.001 | 2.40 (1.89–3.05) | <0.001 | 1.53 (1.30–1.81) | <0.001 |
| AUC  (full model) | 0.82  (0.82–0.83) |  | 0.73  (0.72–0.74) |  | 0.74  (0.71–0.77) |  | 0.63  (0.61–0.65) |  |

**Supplementary table 19** Association between polygenic risk score for atrial fibrillation and postoperative atrial fibrillation in cardiothoracic and non-cardiothoracic surgery subgroups. Results are shown after adjustment for age and sex

|  | **All patients** |  | **Cardiothoracic surgery** |  | **Non-cardiothoracic surgery** |  |
| --- | --- | --- | --- | --- | --- | --- |
| **Term** | **Odds ratio (95% CI)** | **P value** | **Odds ratio (95% CI)** | **P value** | **Odds ratio (95% CI)** | **P value** |
| PGS quintile 1 | 1.00 (reference) | - | 1.00 (reference) | - | 1.00 (reference) | - |
| PGS quintile 2 | 1.50 (1.28 – 1.77) | <0.001 | 1.46 (1.14 – 1.89) | 0.003 | 1.54 (1.24 – 1.91) | <0.001 |
| PGS quintile 3 | 2.11 (1.81 – 2.46) | <0.001 | 1.95 (1.53 – 2.49) | <0.001 | 2.21 (1.80 – 2.71) | <0.001 |
| PGS quintile 4 | 2.76 (2.39 – 3.21) | <0.001 | 1.95 (1.54 – 2.50) | <0.001 | 3.35 (2.77 – 4.07) | <0.001 |
| PGS quintile 5 | 4.92 (4.28 – 5.67) | <0.001 | 2.64 (2.10 – 3.33) | <0.001 | 6.50 (5.43 – 7.82) | <0.001 |
| Age | 1.20 (1.20 – 1.21) | <0.001 | 1.14 (1.13 – 1.16) | <0.001 | 1.23 (1.22 – 1.25) | <0.001 |
| Sex - male | 2.18 (2.02 – 2.37) | <0.001 | 1.18 (1.02 – 1.38) | 0.03 | 2.04 (1.85 – 2.24) | <0.001 |

**Supplementary table 20** Post hoc power calculation for primary GWAS

The table below shows post hoc power across a range of effect sizes (odds ratios, OR) and minimum allele frequencies (MAF) calculated using the *genpwr* package ([10.32614/CRAN.package.genpwr](https://doi.org/10.32614/CRAN.package.genpwr)). Power varied across clinical outcomes due to differences in sample size and case fraction. For AF, AKI and SSI, the study was well powered to detected effect sizes of OR ≥1.3 at a MAF ≥0.1. Stroke and AMI were underpowered, with AMI failing to reach 80% power up to a MAF of 0.8 and OR of 1.5. The study had negligible power to detect effect sizes at an OR ≤1.1. Green cells = power ≥80%; orange ≥70%.

|  |  | **Odds ratio** | | | | | |
| --- | --- | --- | --- | --- | --- | --- | --- |
| **Outcome** | **MAF** | **1.05** | **1.1** | **1.2** | **1.3** | **1.4** | **1.5** |
| **AF** | **0.01** | 1.80E-07 | 1.10E-06 | 2.63E-05 | 3.70E-04 | 0.003 | 0.02 |
|  | **0.05** | 1.60E-06 | 5.10E-05 | 0.01 | 0.18 | 0.68 | 0.97 |
|  | **0.1** | 6.30E-06 | 4.70E-04 | 0.1 | 0.76 | 1 | 1 |
|  | **0.2** | 3.00E-05 | 4.60E-03 | 0.53 | 1.00 | 1 | 1 |
|  | **0.5** | 1.20E-04 | 0.03 | 0.89 | 1.00 | 1 | 1 |
| **AKI** | **0.01** | 2.00E-07 | 1.30E-06 | 3.45E-05 | 5.20E-04 | 0.005 | 0.03 |
|  | **0.05** | 1.90E-06 | 6.80E-05 | 0.01 | 0.23 | 0.77 | 0.98 |
|  | **0.1** | 8.00E-06 | 6.70E-04 | 0.14 | 0.84 | 1 | 1 |
|  | **0.2** | 4.00E-05 | 6.60E-03 | 0.63 | 1.00 | 1 | 1 |
|  | **0.5** | 1.70E-04 | 0.04 | 0.94 | 1.00 | 1 | 1 |
| **AMI** | **0.01** | 6.00E-08 | 9.40E-08 | 3.00E-07 | 9.00E-07 | 2.00E-06 | 1.00E-05 |
|  | **0.05** | 1.10E-07 | 3.70E-07 | 4.20E-06 | 3.37E-05 | 2.00E-04 | 0.001 |
|  | **0.1** | 1.70E-07 | 9.90E-07 | 2.15E-05 | 2.78E-04 | 0.002 | 0.01 |
|  | **0.2** | 3.10E-07 | 2.90E-06 | 1.26E-04 | 2.28E-03 | 0.02 | 0.1 |
|  | **0.5** | 5.50E-07 | 7.70E-06 | 4.95E-04 | 9.31E-03 | 0.07 | 0.24 |
| **SSI** | **0.01** | 1.80E-07 | 1.10E-06 | 2.53E-05 | 3.52E-04 | 0 | 0.02 |
|  | **0.05** | 1.50E-06 | 4.90E-05 | 0.01 | 0.17 | 0.66 | 0.96 |
|  | **0.1** | 6.10E-06 | 4.50E-04 | 0.1 | 0.75 | 0.99 | 1 |
|  | **0.2** | 2.90E-05 | 4.30E-03 | 0.52 | 0.99 | 1 | 1 |
|  | **0.5** | 1.20E-04 | 0.02 | 0.88 | 1.00 | 1 | 1 |
| **Stroke** | **0.01** | 7.20E-08 | 1.50E-07 | 1.00E-06 | 3.50E-06 | 1.00E-05 | 5.00E-05 |
|  | **0.05** | 1.80E-07 | 1.10E-06 | 3.00E-05 | 3.75E-04 | 0.003 | 0.02 |
|  | **0.1** | 3.70E-07 | 4.00E-06 | 2.00E-04 | 0.004 | 0.04 | 0.18 |
|  | **0.2** | 8.40E-07 | 1.70E-05 | 0.002 | 0.04 | 0.26 | 0.67 |
|  | **0.5** | 1.80E-06 | 6.00E-05 | 0.01 | 0.15 | 0.57 | 0.9 |

**Supplementary Figures**

1. **Manhattan plot – acute kidney injury**
2. **Q-Q plot – acute kidney injury. Genomic inflation factor (λ) 1.04**
3. **Manhattan plot – acute myocardial infarction**
4. **Q-Q plot – acute myocardial infarction. Genomic inflation factor (λ) 1.02**
5. **Manhattan plot – stroke**
6. **Q-Q plot – stroke. Genomic inflation factor (λ) 1.00**
7. **Manhattan plot – surgical site infection**
8. **Q-Q plot – surgical site infection. Genomic inflation factor (λ) 1.01**
9. **Stratified analysis of SNP effects across surgical specialties for postoperative atrial fibrillation**
10. **Stratified analysis of SNP effects across surgical specialties for postoperative acute kidney injury**
11. **Stratified analysis of SNP effects across surgical specialties for postoperative acute myocardial infarction**
12. **Stratified analysis of SNP effects across surgical specialties for postoperative stroke**

**Supplementary figure 1** Manhattan plot – acute kidney injury

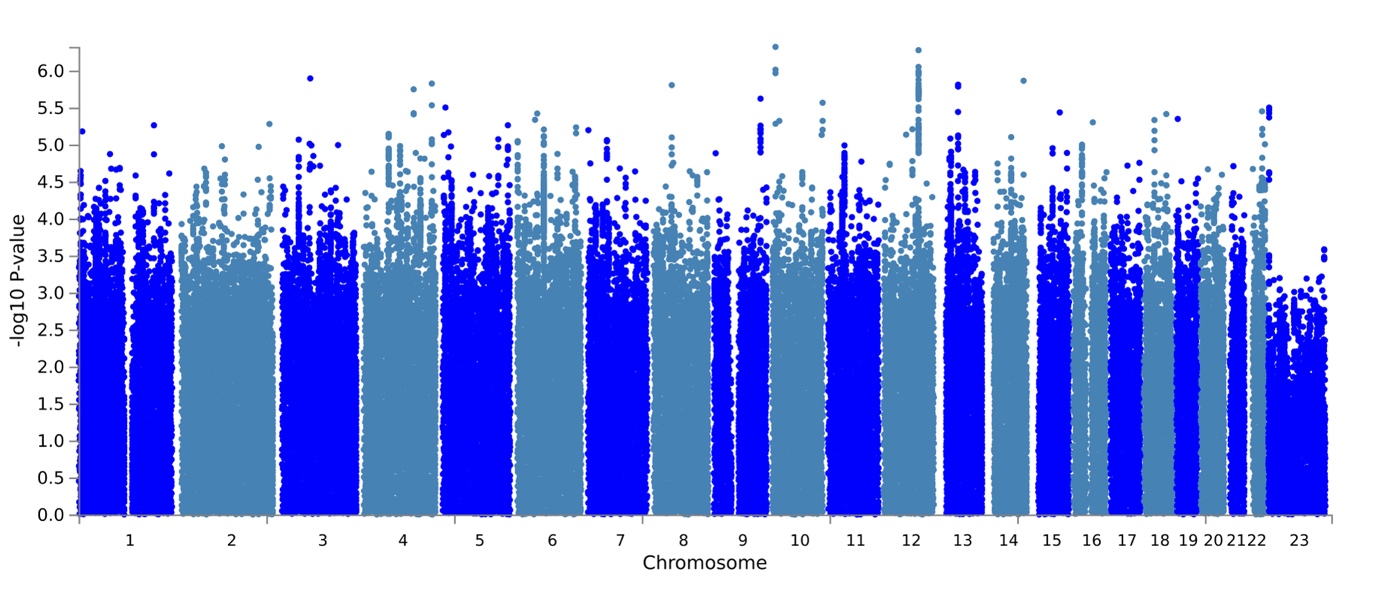

**Supplementary figure 2** Q-Q plot – acute kidney injury. Genomic inflation factor (λ) 1.04

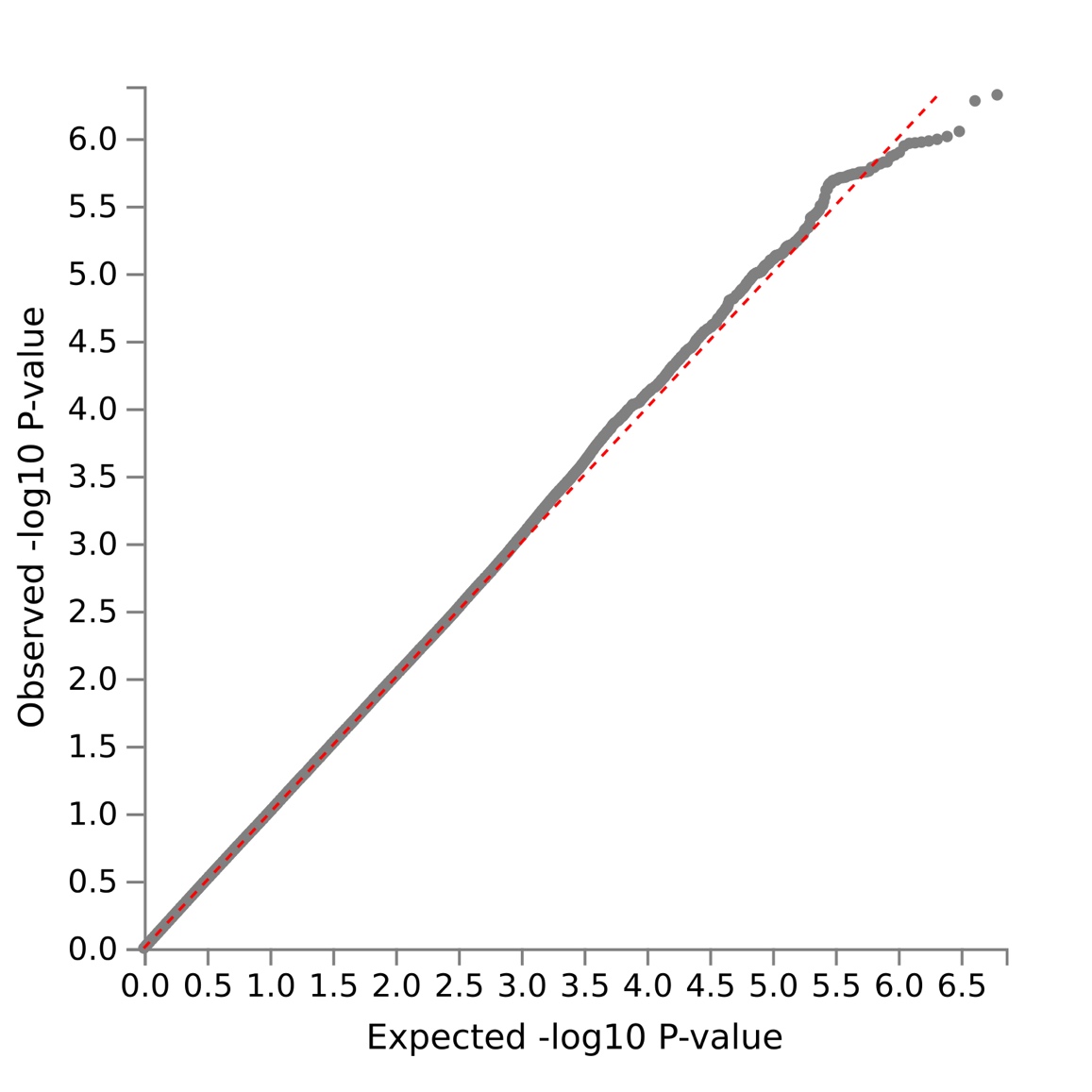

**Supplementary figure 3** Manhattan plot – acute myocardial infarction

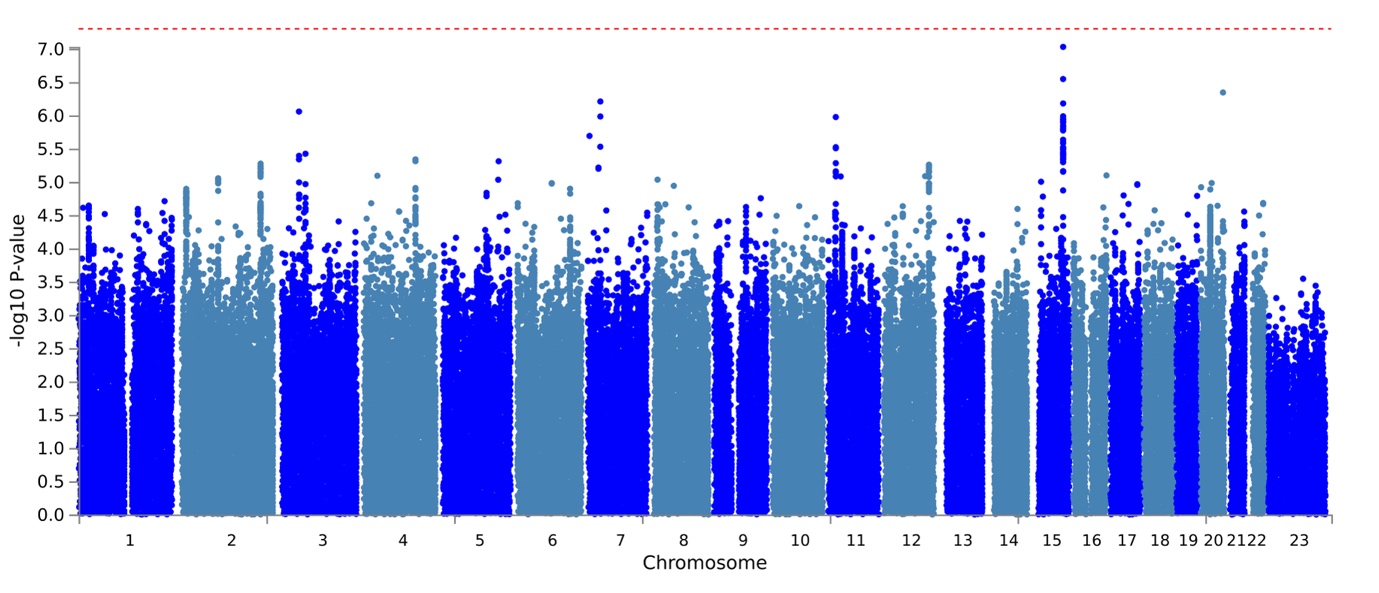

**Supplementary figure 4** Q-Q plot – acute myocardial infarction. Genomic inflation factor (λ) 1.02

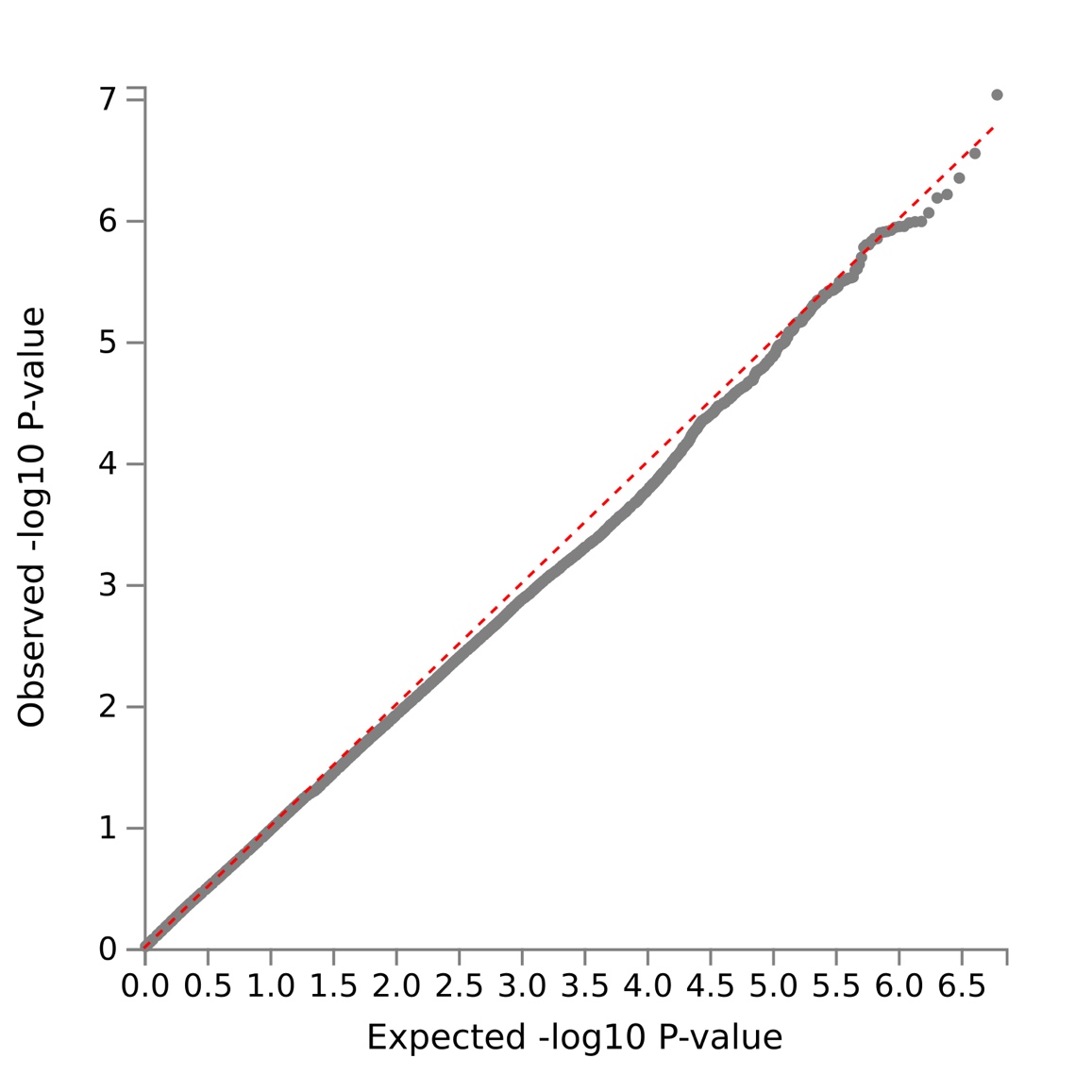

**Supplementary figure 5** Manhattan plot – stroke

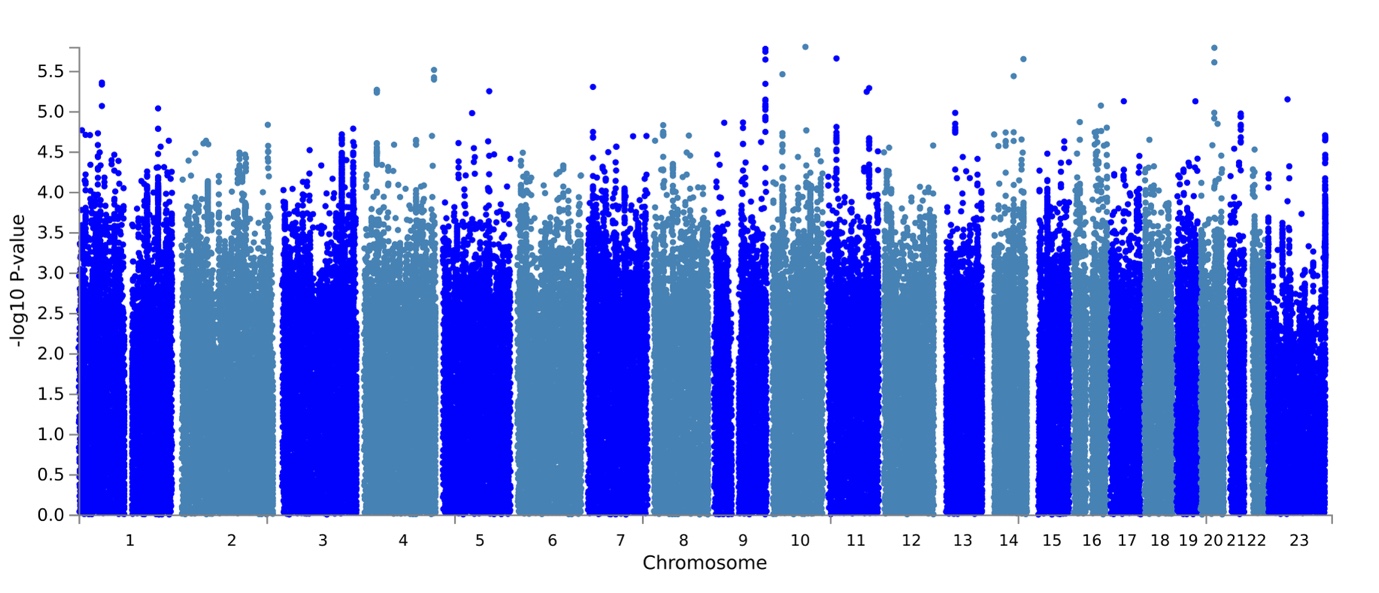

**Supplementary figure 6** Q-Q plot – stroke. Genomic inflation factor (λ) 1.00

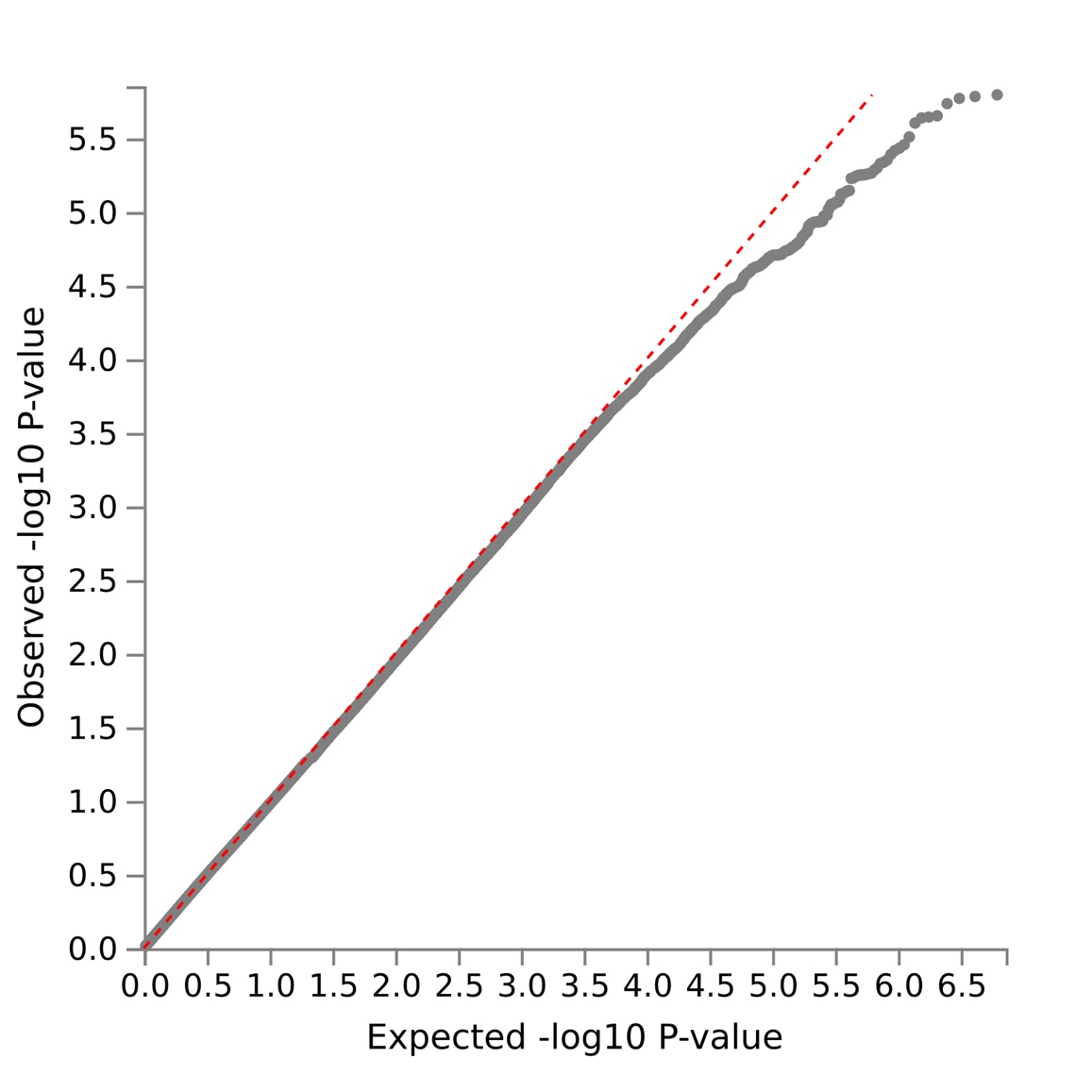

**Supplementary figure 7** Manhattan plot – surgical site infection

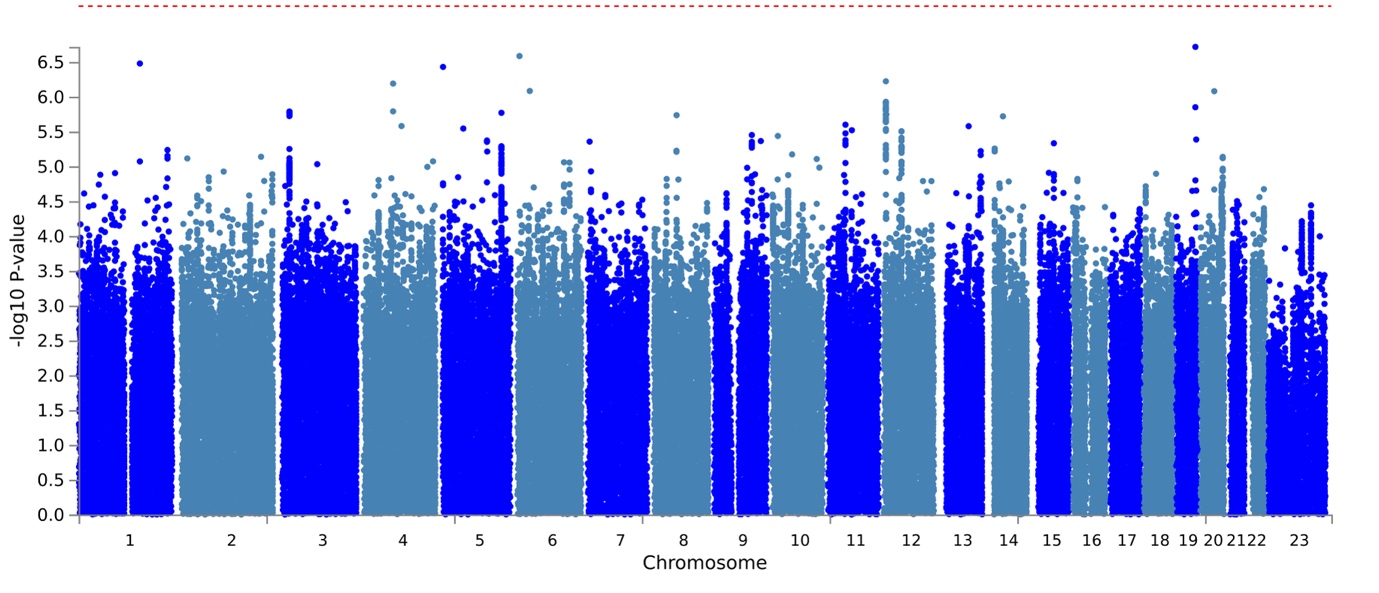

**Supplementary figure 8** Q-Q plot – surgical site infection. Genomic inflation factor (λ) 1.01

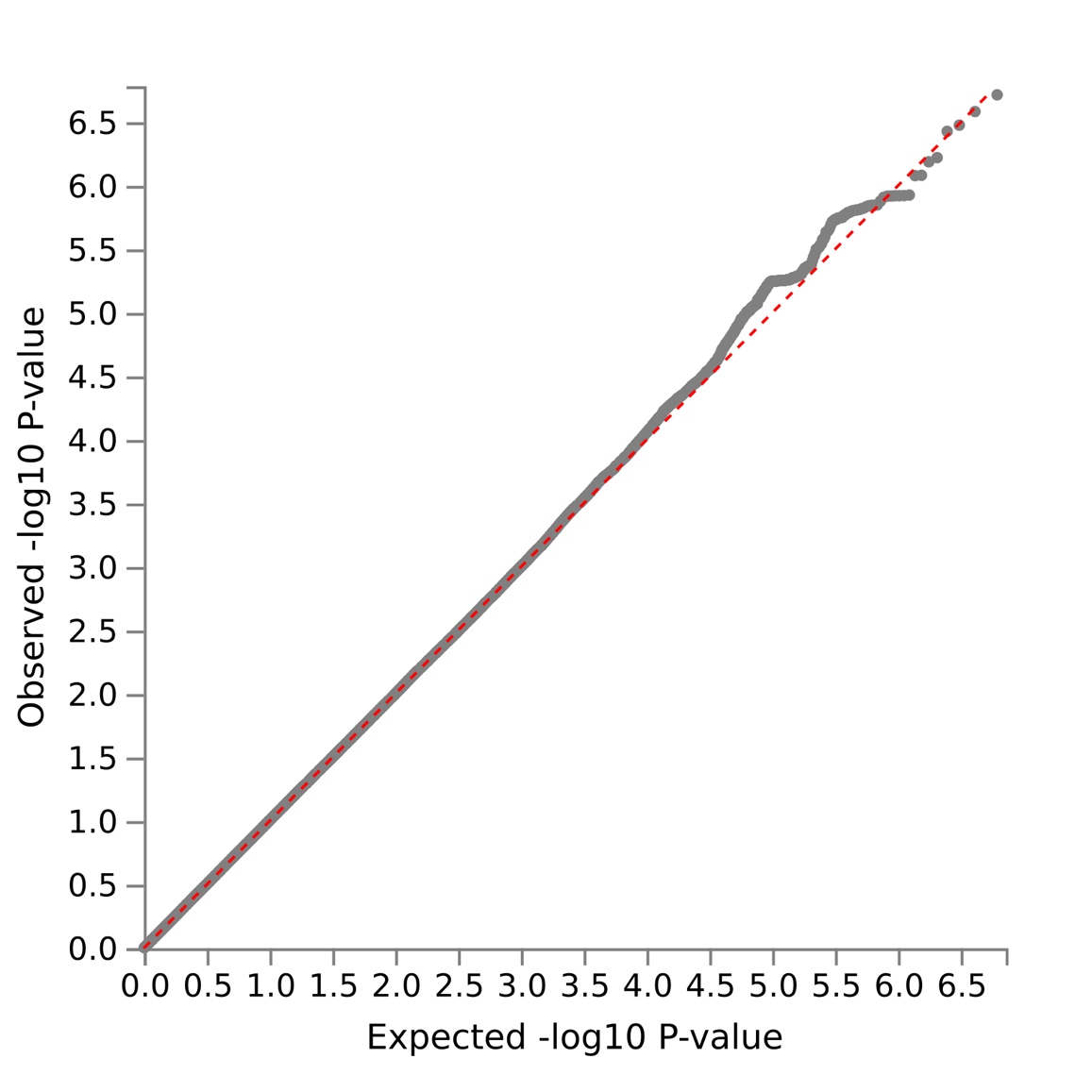

**Supplementary figure 9** Stratified analysis of SNP effects across surgical specialties for postoperative atrial fibrillation. Markers are odds ratios with 95% confidence interval error bars. X-axis limited to 3.0, error bars beyond this not shown.

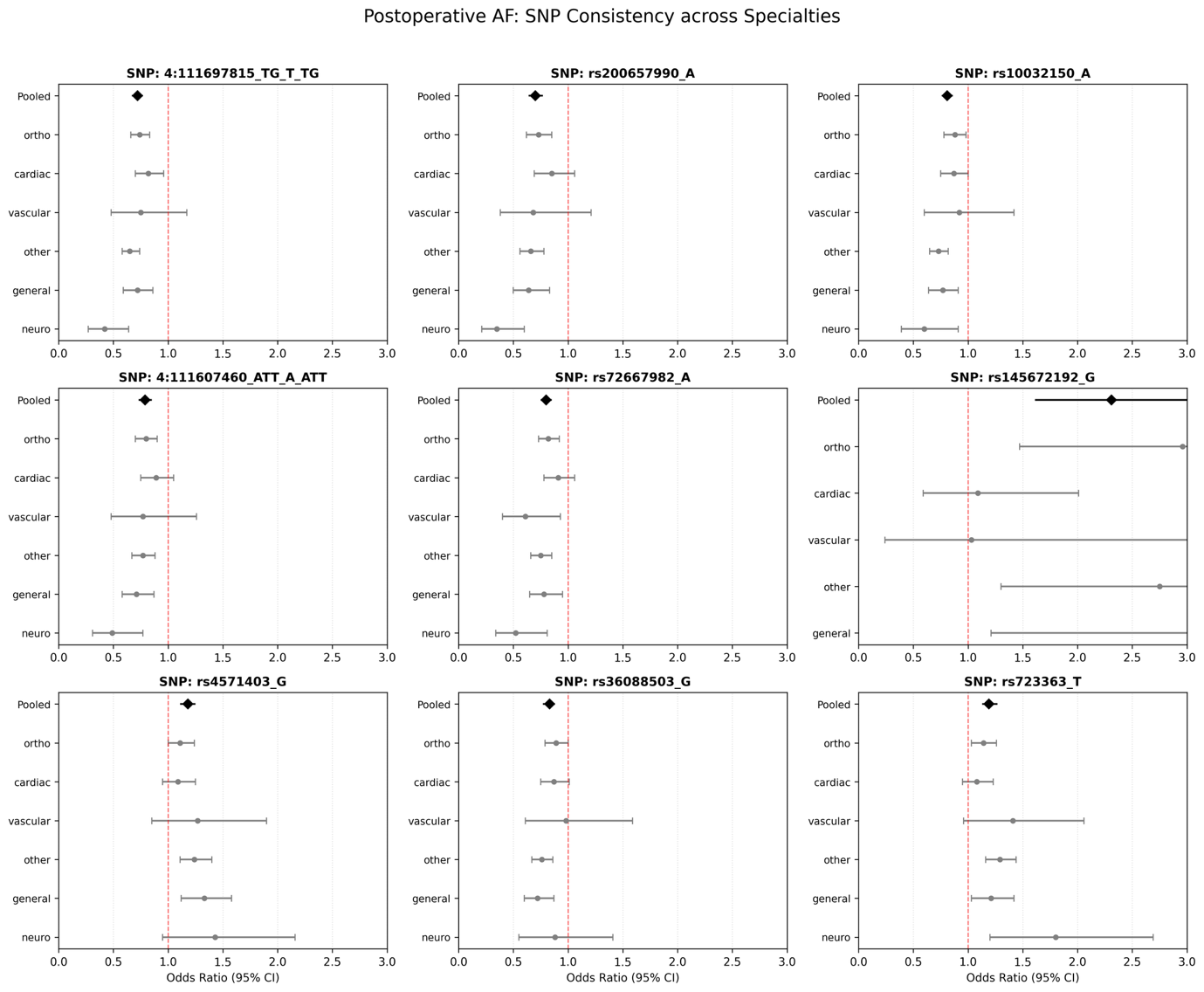

**Supplementary figure 10** Stratified analysis of SNP effects across surgical specialties for postoperative acute kidney injury. Markers are odds ratios with 95% confidence interval error bars.

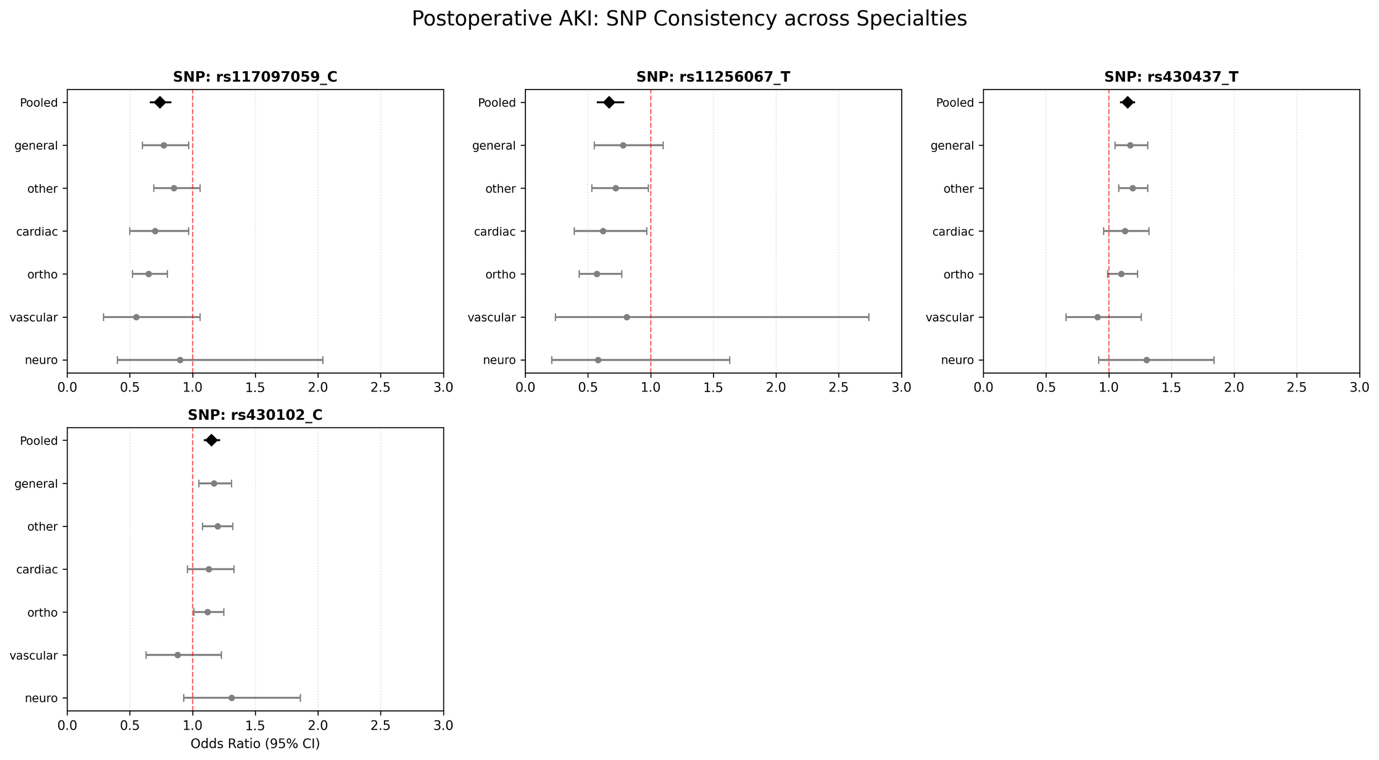

**Supplementary figure 11** Stratified analysis of SNP effects across surgical specialties for postoperative acute myocardial infarction. Markers are odds ratios with 95% confidence interval error bars. X-axis limited to 3.0, error bars beyond this not shown.

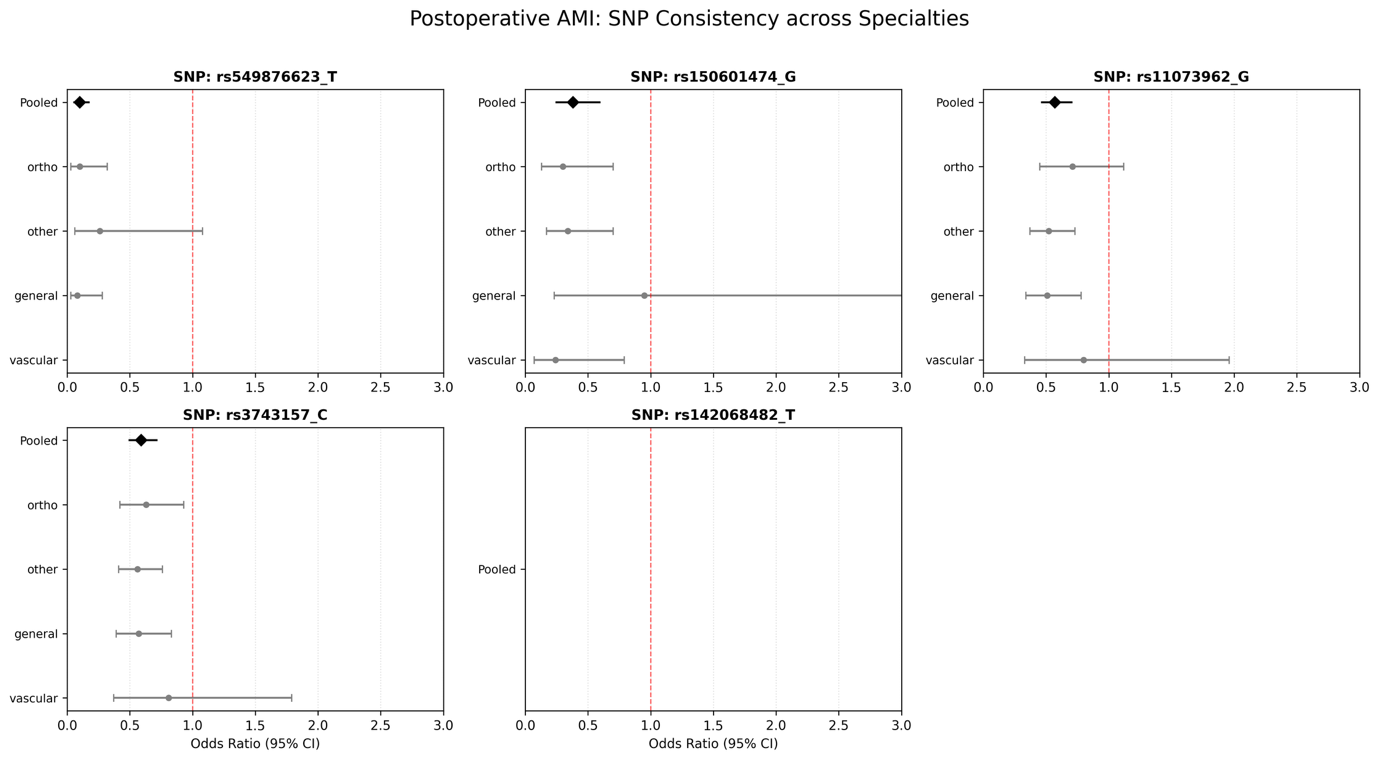

**Supplementary figure 12** Stratified analysis of SNP effects across surgical specialties for postoperative surgical site infection. Markers are odds ratios with 95% confidence interval error bars. X-axis limited to 3.0, error bars beyond this not shown.

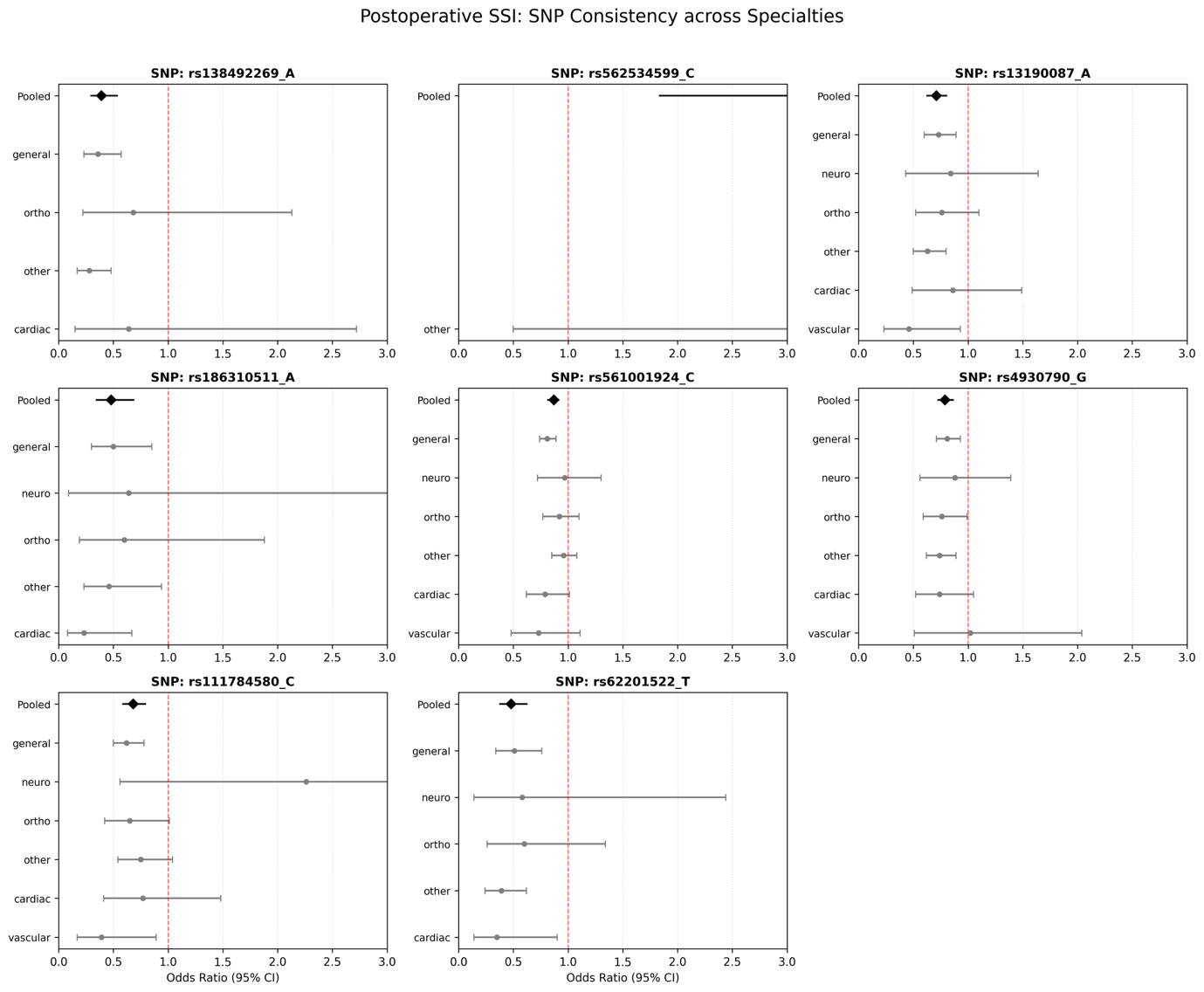
